# Molecular Landscape and Contemporary Prognostic Signatures of Gliomas

**DOI:** 10.1101/2023.09.09.23295096

**Authors:** Hia S. Ghosh, Ruchit V. Patel, Eleanor Woodward, Noah F. Greenwald, Varun M. Bhave, Eduardo A. Maury, Gregory Cello, Samantha E. Hoffman, Yvonne Li, Hersh Gupta, Liam F. Spurr, Jayne Vogelzang, Mehdi Touat, Frank Dubois, Andrew D. Cherniack, Xiaopeng Guo, Sherwin Tavakol, Gino Cioffi, Neal I. Lindeman, Azra H. Ligon, E. Antonio Chiocca, David A. Reardon, Patrick Y. Wen, David Meredith, Sandro Santagata, Jill S. Barnholtz-Sloan, Keith L. Ligon, Rameen Beroukhim, Wenya Linda Bi

## Abstract

Molecularly-driven treatments are expanding options for patients with gliomas, driving a need for molecularly-informed prognostic information. To characterize the genomic landscape and contemporary outcomes of gliomas, we analyzed 4,400 gliomas from multi-institutional datasets and The Cancer Genome Atlas (TCGA): 2,195 glioblastoma, 1,198 *IDH1/2*-mutant astrocytoma, 531 oligodendroglioma, 271 other *IDH1/2*-wildtype glioma, and 205 pediatric-type glioma. Molecular classification updated 27.4% of gliomas from their original histopathologic diagnosis. Canonical alterations helped categorize glioma subtypes, revealing mutually exclusive alterations within tumorigenic pathways. Across each glioma subtype, non-TCGA patients had longer survival compared to TCGA patients. Several novel prognostic alterations emerged, including *NF1* alteration and 21q loss in glioblastoma, and *EGFR* amplification and 22q loss in *IDH1/2-*mutant astrocytoma. Certain prognostic features varied across age, with decreasing prevalence of *IDH1/2*-mutation over time while *MGMT*-methylation remained steady. Our findings provide a framework for further exploration and validation of glioma prognostic indicators in clinically representative cohorts and trials.

## Introduction

The classification of gliomas, the most common malignant brain tumor in adults, has undergone significant transformation with routine incorporation of molecular markers, which improve prediction of tumor behavior, response to therapy, and patient outcomes.^1–6^ These molecular features were initially elucidated through large-scale efforts, including The Cancer Genome Atlas (TCGA), which were conducted before changes in treatment standards of care and the introduction of current classification schemes.^7–9^ Subsequent large-scale investigations have probed the molecular underpinnings of specific glioma subtypes or developed datasets that focus on particular molecular or clinical features.^10–15^ In our study, we aimed to comprehensively characterize the genetic landscape of gliomas, evaluate trends in patient survival, and identify prognostic genomic alterations using a large, multi-institutional dataset of molecularly annotated gliomas, which represents a contemporary and clinically heterogeneous cohort. Our integrated analysis demonstrates distinct mutational profiles across different glioma subtypes and patient lifespan, significant clinical and molecular differences between contemporary patient cohorts and historical ones, and subtype-dependent characteristics that are predictors of overall survival.

## Results

### Molecular Features Refine Histopathological Diagnoses

We identified 4,400 unique patients (median age 52 years, range 0-94 years) with molecularly annotated gliomas from three datasets: DFCI/BWH (n=1565), GENIE (n=2063), and TCGA (n=772; Fig. 1A; Table 1). This spanned 2,195 glioblastoma, 1,198 *IDH1/2*-mutant astrocytoma, 531 *IDH1/2*-mutant oligodendroglioma, 271 other *IDH1/2-*wildtype glioma, and 205 pediatric-type glioma (89 low-grade, 116 high-grade), all classified according to the World Health Organization (WHO) Classification of Tumors of the Central Nervous System 2021 guidelines and the six cIMPACT-NOW Updates.^5,6^

**Figure 1:**
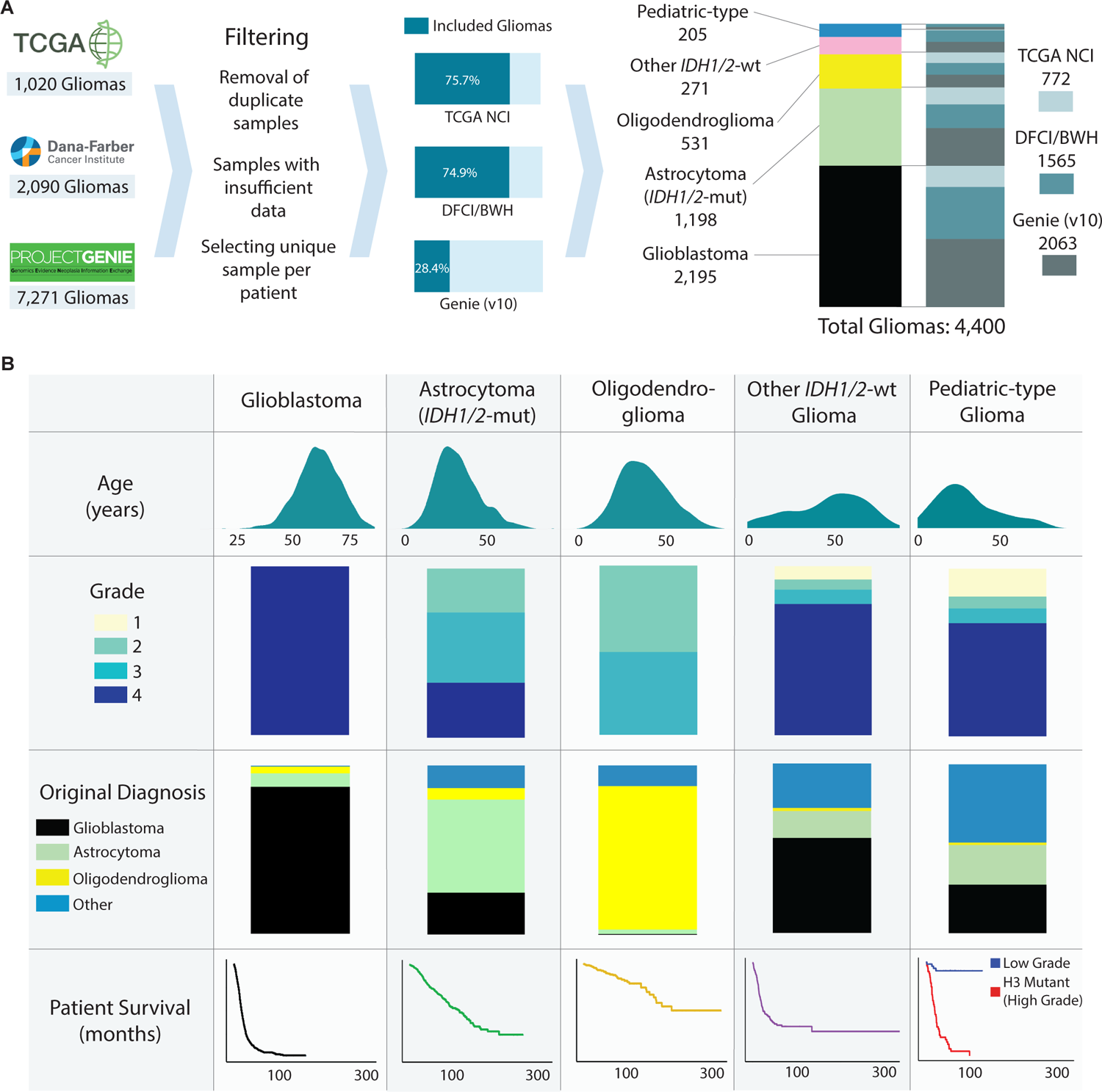
Cohort overview. (A) Overview of constructing a pooled molecularly annotated glioma cohort. (B) Summary of the pooled glioma cohort.

Molecular classification significantly refined glioma subtypes from their original histopathological diagnoses. Among the molecularly classified glioblastomas, 87.4% were consistent with their original designation, while 8.0% were updated in classification as varying grades of astrocytomas, 4.0% as other gliomas, and 0.6% as oligodendrogliomas (Fig. 1B, Table 1). 7.4% of glioblastoma were upgraded from grades 2/3 to grade 4 after molecular classification. Molecularly defined *IDH1/2*-mutant astrocytomas (grades 2-4) showed the greatest heterogeneity in their original histopathologic classifications–55.0% were previously classified as astrocytomas, 24.8% as glioblastoma, 13.4% as other gliomas, and 6.8% as oligodendrogliomas. By contrast, *IDH1/2*-mutant 1p/19q co-deleted oligodendrogliomas showed higher concordance with their histopathologic designation, with 84.8% aligning with their original classification as oligodendrogliomas, 12.2% as other gliomas, 2.6% as astrocytomas, and 0.4% as glioblastomas. Pediatric-type gliomas were enriched in glioblastoma (28.8%) and astrocytoma (23.4%) while other *IDH1/2*-wildtype gliomas were largely histologically characterized as glioblastoma (56.1%).

### Molecular Alterations Vary Across Glioma Subtypes

Canonical molecular alterations categorized the dominant glioma subtypes (Table 1, Fig. 2A, Supplement 1). Glioblastoma most frequently harbored homozygous deletion of *CDKN2A/B* (56.5%), *EGFR* amplification (47.5%) and *PTEN* alterations (47.7%) compared to other subtypes, and a majority (91.0%) also carried *TERT* promoter alterations. The majority of glioblastoma had whole chromosome 7 gain/chromosome 10 loss (7+/10-) (57.7%), while partial 7+/10-alterations were found in an additional 31.1% of glioblastomas. When whole 7+/10-were observed in non-glioblastoma subtypes, it was exclusively present in grade 3 and 4 tumors. *IDH1/2*-mutant astrocytomas frequently had *TP53* (91.7%) and *ATRX* (61.3%) alterations, which were relatively less common in glioblastoma and rare in oligodendroglioma. *EGFR* amplifications were rare in *IDH1/2-*mutant astrocytomas (1.7%), and they were exclusive to high grade tumors. Oligodendrogliomas had frequent alterations in *TERT* promoter (94.3%) and *CIC* (69.1%) as well as a low prevalence of *TP53* (7.5%) and *ATRX* (6.4%) alterations. Low-grade pediatric-type gliomas were enriched with *BRAF* mutations and rearrangements (39.3%, 25.8% respectively) and *FGFR1* alterations (30.3%), while *TP53* alterations (39.9%) and homozygous deletion of *CDKN2A/B* (29.9%) were prevalent in other *IDH1/2*-wildtype gliomas.

**Figure 2:**
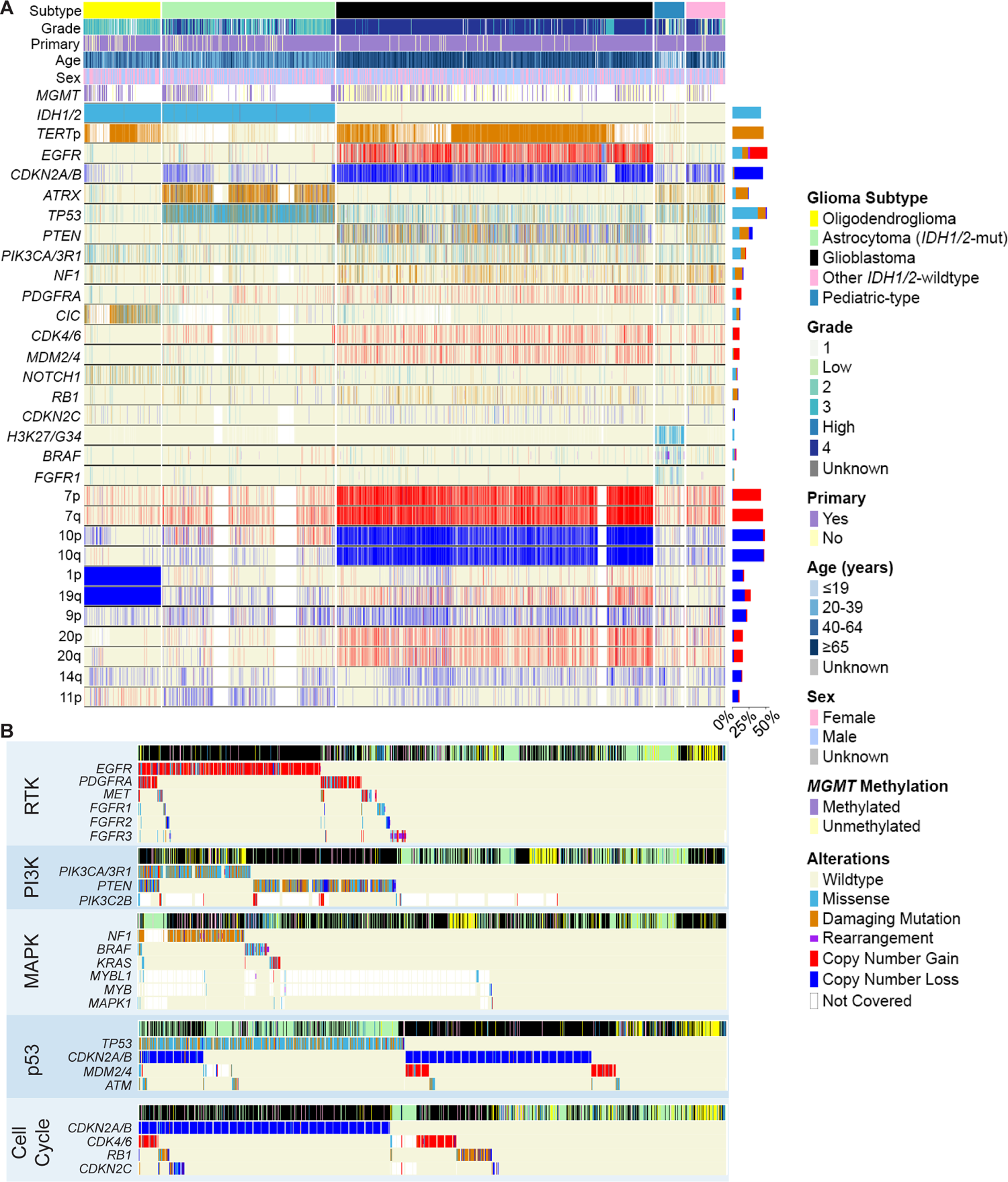
Mutational signatures in glioma. (A) Mutational signatures of gliomas stratified by tumor subtype. Genes were filtered out of the co-mutation plot if alteration prevalence was low or if not involved in molecular glioma classification guidelines. (B) Molecular alterations within the same tumorigenic pathway were frequently mutually exclusive in glioma.

We next assessed five frequently altered pathways associated with tumorigenesis: receptor tyrosine kinase (RTK), phosphoinositide-3-kinase (PI3K), mitogen-activated protein kinase (MAPK), p53, and cell cycle. Gliomas frequently exhibited concurrent aberrations in these pathways, with glioblastoma having an average of 2.8 pathways affected (median: 3), *IDH1/2*-mutant astrocytoma with 1.7 (median: 1), and oligodendroglioma with 1.5 (median: 1, Supplement 2). However, gliomas rarely harbored multiple alterations within each pathway, a phenomenon commonly observed in cancer genomes (Fig. 2B).^16^ Amongst RTKs, alterations in *EGFR* showed limited co-occurrence with *PDGFRA* (8.4%), *MET* (4.6%), and *FGFR1-3* (2.7%, 2.4%, 2.4%). Of all RTKs analyzed, *EGFR* had the highest prevalence of rearrangement. In the PI3K pathway, there was minimal co-occurrence of *PIK3CA/3R1* and *PTEN* mutations (8.3%), especially in glioblastoma. Across the MAPK pathway, mutations in *NF1*, *BRAF*, and *KRAS* were almost entirely mutually exclusive: mutations in *NF1* co-occurred with *BRAF* in 3.6% and with *KRAS* in 2.2% of altered cases, while *BRAF* and *KRAS* mutations co-occurred in 1.3% of altered cases. In the p53 pathway, while *TP53* and *CDKN2A/B* alterations overlapped in a subset of glioblastomas and *IDH1/2*-mutant astrocytomas, *TP53* mutation predominated in *IDH1/2*-mutant astrocytomas, while *CDKN2A/B* alterations predominated in glioblastomas. Additionally, focal amplifications of *MDM2 or MDM4*, known regulators of p53, were also seen in a subpopulation of glioblastomas without *TP53* or *CDKN2A/B* alteration. Finally, amongst other cell cycle mediators, there was minimal overlap between alterations in *CDKN2A/B*, *CDK4/6*, *RB1*, and *CDKN2C*.

### Genomic Correlates and Distance Distinguish Glioma Subtypes

When we stratified the glioma subtypes, we observed distinct relationships between clinical variables and molecular alterations (Fig. 3A-C). In glioblastoma, alterations in *CDKN2A/B* and *PDGFRA* alterations were significantly enriched in patients ≥65 years old while there was a depletion of these alterations in patients between 40-64 years old (Fig. 3A). *RB1* alterations showed the inverse relationship, with enrichment in patients between 40-64 years old. Age also showed associations with molecular alterations in *IDH1/2*-mutant astrocytomas and oligodendrogliomas, where patients <40 years old exhibited distinct correlated molecular alterations compared to those between 40-64 and ≥65 years (Fig. 3B-C). Moreover, there was a clear genomic distinction across different grades within *IDH1/2-*mutant astrocytomas and oligodendrogliomas. Grade 4 *IDH1/2*-mutant astrocytomas and grade 3 oligodendrogliomas showed positive correlations with a range of molecular alterations, while grades 2/3 in *IDH1/2*-mutant astrocytomas and grade 2 oligodendrogliomas were negatively correlated with nearly all of the same molecular alterations.

**Figure 3:**
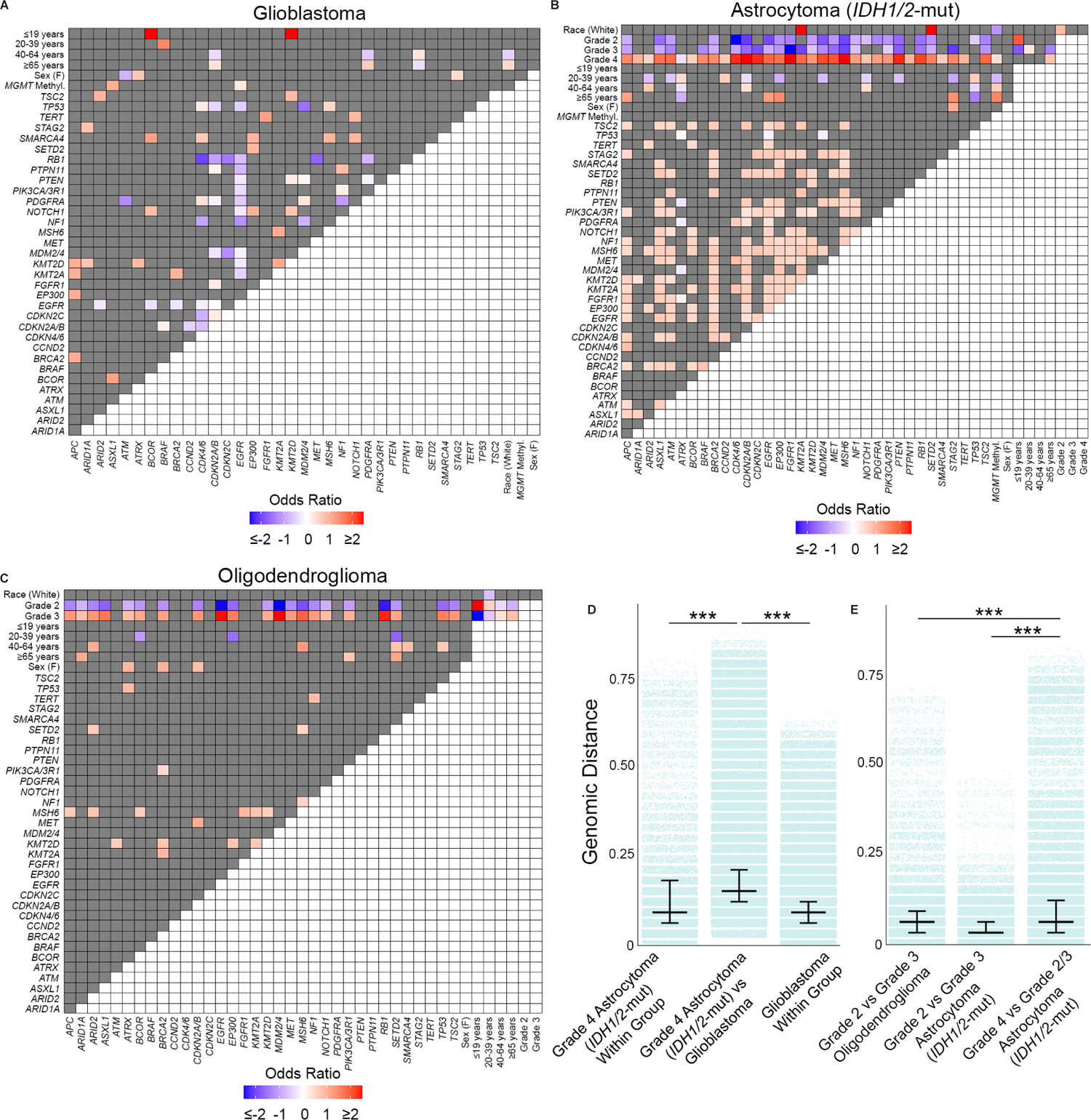
Genomic correlations in glioma subtypes. Heatmap of positively and inversely correlated genes for (A) glioblastoma, (B) *IDH1/*2-mutant astrocytoma, and (C) oligodendroglioma. (D) The genomic distance between grade 4 *IDH1/2-*mutant astrocytomas and glioblastoma was greater than the genomic distance within each glioma group. (E) Grade 4 *IDH1/2*-mutant astrocytoma were more genomically distinct from Grade 2/3 *IDH1/2*-mutant astrocytoma, with greater genomic separation compared to grade 2 and grade 3 *IDH1/2*-mutant astrocytoma or between grade 2 and grade 3 oligodendroglioma. p<0.001 (***)

Relationships between molecular alterations also emerged. In glioblastoma, *EGFR* alterations were inversely correlated with a wide range of molecular alterations, including *TP53*. Compared to glioblastoma or oligodendroglioma, *IDH1/2*-mutant astrocytomas appeared more molecularly heterogenous. Amongst *IDH1/2*-mutant astrocytomas, *CDKN2A/B* and *EGFR* alterations positively correlated with several deleterious alterations across RTK, PI3K, MAPK pathways. Notably, *EGFR* alterations were positively correlated with both *CDKN2A/B* and *PDGFRA* alterations, reflecting the co-occurrence of canonical glioblastoma molecular alterations in *IDH1/2*-mutant astrocytoma.

We quantified the heterogeneity of glioma genomes using genomic distance (Supplement 3). We observed greater genomic variability between gliomas of different subtypes than within a subtype. Specifically, the genomic difference between glioblastoma and *IDH1/2*-mutant astrocytoma (median Jaccard Distance (JD): 0.147) was significantly higher than the genomic heterogeneity within glioblastoma (median JD: 0.088, p<0.001) or *IDH1/2*-mutant astrocytoma (median JD: 0.059, p<0.001). Similarly, the genomic distance between glioblastoma and oligodendroglioma (median JD: 0.118) and between *IDH1/2-*mutant astrocytoma and oligodendroglioma (median JD: 0.088) was greater than the genomic heterogeneity within each respective glioma subtype (all p<0.001).

When we subdivided gliomas by grade, we saw distinct patterns of genomic distance. Grade 4 *IDH1/2*-mutant astrocytomas and glioblastomas were more genomically distant from each other (median JD: 0.147) compared to genomic differences within grade 4 *IDH1/2*-mutant astrocytomas (median JD: 0.088) or within glioblastomas (median JD: 0.088, p<0.001, Fig. 3D). Furthermore, grade 4 *IDH1/2*-mutant astrocytomas showed a greater genomic distance from grade 2/3 *IDH1/2-*mutant astrocytomas than between grade 2 and 3 *IDH1/2*-mutant astrocytomas (p<0.001, Fig. 3D). This distance between grade 4 versus grade 2-3 *IDH1/2*-mutant astrocytoma outstripped the genomic distance between grade 3 versus grade 2 oligodendrogliomas (p<0.001, Fig. 3D). This demonstrated the unique genomic makeup of grade 4 *IDH1/2*-mutant astrocytomas, distinguishing them from glioblastoma and other grade 2 and 3 *IDH1/2*-mutant gliomas.

### Glioma Cohort and Subtype Influence Survival

Among patients ≥20 years old with primary gliomas, the median survival varied across glioblastoma (18.0 months, range: 0.1-164.0 months), *IDH1/2-*mutant astrocytoma (118.5 months, range: 0.1-262.2 months), and oligodendroglioma (213.9 months, range: 0.1-324.4 months). Notably, there was a significant difference in median survival between patients in the non-TCGA cohort and those in the TCGA cohort. For glioblastoma, non-TCGA patients had a median overall survival of 19.0 months (range: 0.2-164.0 months), which was 26.7% longer than the median overall survival of TCGA patients (15.0 months, range: 0.1-95.3 months, p<0.001, Fig. 4A). The difference in median survival between non-TCGA and TCGA cohorts was even more pronounced for *IDH1/2-*mutant astrocytoma. Non-TCGA patients had a median survival of 136.8 months (range: 1.0-262.2 months), 55.6% longer than the median survival of TCGA patients at 87.9 months (range: 0.1-157.1 months, p=0.0002, Fig. 4C). Similarly, in the case of oligodendroglioma, non-TCGA patients had a median survival of 307.5 months (range: 0.3-324.4 months), 127.8% longer than the median survival of TCGA patients at 135.0 months (range: 0.1-183.3 months, p<0.0001, Fig. 4E).

**Figure 4:**
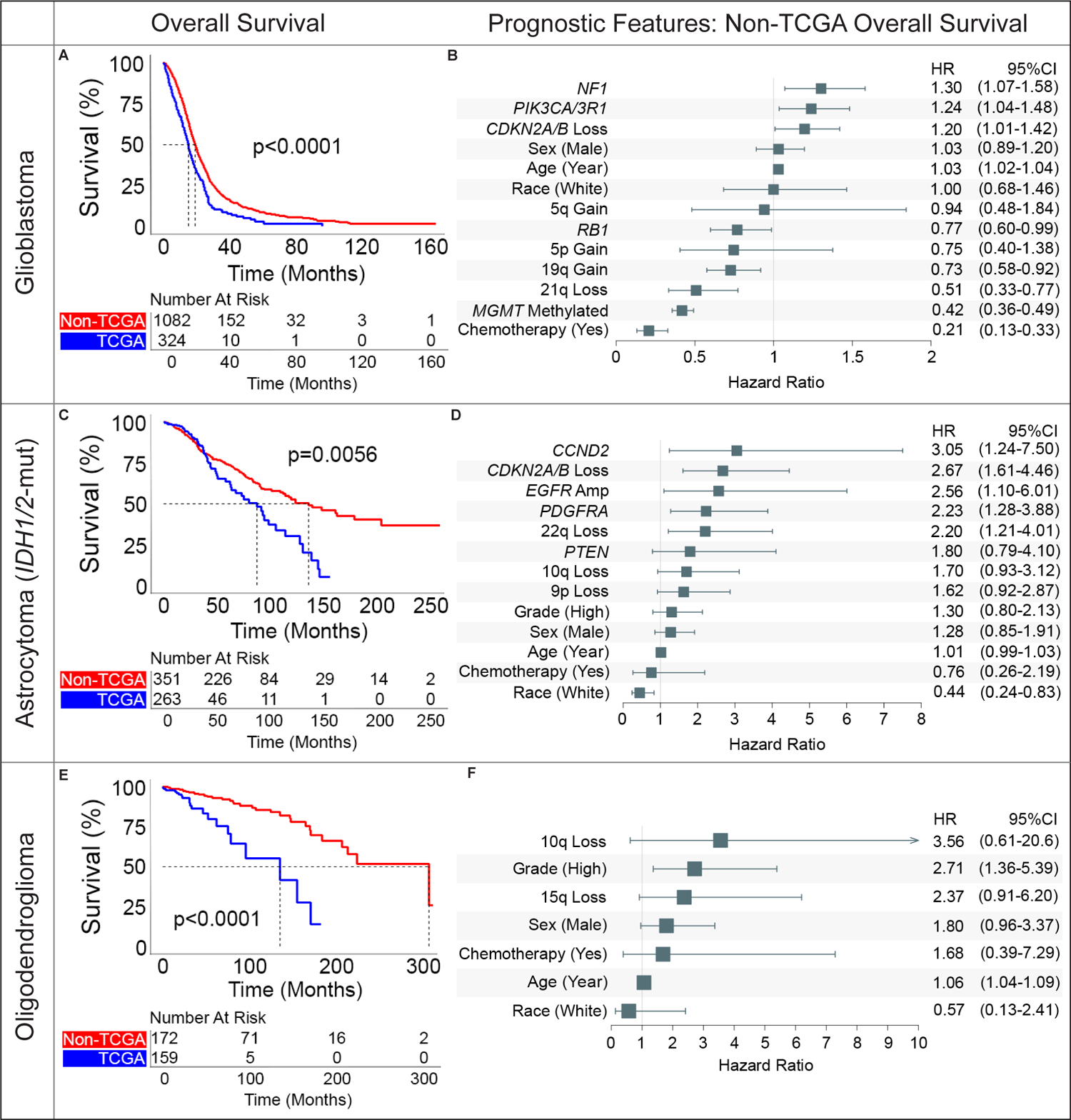
Overall survival and prognostic features in glioma subtypes. Kaplan-Meier curves demonstrate overall survival in the non-TCGA cohort exceed that of the TCGA cohort for patients with newly diagnosed (A) glioblastoma, (C) *IDH1/2*-mutant astrocytoma, and (E) oligodendroglioma. Multivariate adjusted clinical and molecular features predictive of overall patient survival for newly diagnosed (B) glioblastoma, (D) *IDH1/2-*mutant astrocytoma, and (F) oligodendroglioma.

Considering the cohort differences, we analyzed the effect of glioma grade in the non-TCGA cohort to gain an updated view on overall survival across different glioma subtypes. In oligodendroglioma, patients with grade 2 tumors did not reach median survival (range: 0.3-324.4 months) whereas grade 3 oligodendrogliomas had a median survival of 214.0 months (range: 4.7-307.5 months, p=0.001, Supplement 4A). Median survival of grade 2/3 *IDH1/2-*mutant astrocytomas more than doubled that of grade 4 *IDH1/2-*mutant astrocytomas (grade 2: 180.3 months, range: 1.0-262.2 months; grade 3: not reached, range: 2.0-250.3 months; grade 4: 55.9 months, range: 1.8-203.9 months, p<0.0001, Supplement 4B). Non-glioblastoma *IDH1/2*-wildtype gliomas had far more heterogenous survival, with low-grade pediatric-type gliomas showing significantly greater survival compared high-grade pediatric-type gliomas and other *IDH1/2*-wildtype gliomas (Supplement 4C). These updated survival analyses provide further insights into the effect of glioma grade on overall survival within specific subtypes.

### Prognostic Features Vary Across Glioma Subtypes

We identified new and canonical prognostic features within different glioma subtypes after multivariate adjustment in the non-TCGA cohort (Fig. 4). Given the differences in patient survival between non-TCGA and TCGA cohorts, we reassessed the features based on cohort status as a covariate in the analysis.

Across glioblastoma, *IDH1/2*-mutant astrocytoma, and oligodendroglioma, non-TCGA cohort status remained positively prognostic (Supplement 6B, 6D, 6E). Most patients in both cohorts received chemotherapy: 94.8% of patients in non-TCGA cohorts received chemotherapy, while 87.9% of patients in TCGA were labeled as receiving chemotherapy.

In non-TCGA patients with glioblastoma, receipt of chemotherapy (HR: 0.21), methylated *MGMT* (HR: 0.42), 21q loss (HR: 0.51), 19q gain (HR: 0.73), and *RB1* alteration (HR: 0.77) positively affected survival. *NF1* alteration (HR: 1.30), *PIK3CA/3R1* alteration (HR: 1.24), *CDKN2A/B* loss (homozygous or heterozygous loss, HR: 1.20), and increasing age per year (HR: 1.03) negatively impacted survival for glioblastoma (Fig. 4B). Homozygous and heterozygous loss of *CKDN2A/B* were assessed together as both exerted a similar negative effect on overall survival (Supplement 5A). Chromosome 21q loss emerged as a novel positive prognostic feature, with overall survival diverging around the two year mark (Supplement 5B). The prognostic significance of these features differed when assessed in the TCGA cohort and when all cohorts were pooled (Supplement 6A-6B). For example, in the TCGA cohort, features like 21q loss, *NF1* alterations, and *RB1* alterations were not significant. However, male sex emerged as a negative prognostic feature only in the TCGA cohort (HR: 2.07) but was not significant after cohort status was considered. *MGMT*-methylation status trended toward significance in the TCGA cohort (HR: 0.83), likely because methylation status significantly improved survival only when chemotherapy was administered (Supplement 7).

In non-TCGA *IDH1/2-*mutant astrocytomas, several molecular features were identified as negatively prognostic for survival: *CCND2* alteration (HR: 3.05), *CDKN2A/B* loss (homozygous or heterozygous loss, HR: 2.67), *EGFR* amplification (HR: 2.56), *PDGFRA* alteration (HR: 2.23), and 22q loss (HR: 2.20) (Fig. 4D). 10q loss (HR: 1.70, p=0.08) and 9p loss (HR: 1.62, p=0.09) trended toward significance as negative prognostic features. Many of these molecular features only emerged in the non-TCGA cohort and were prognostically heterogenous in the TCGA cohort (Supplement 6C). Consistent with prior results, tumor grade and *MGMT*-methylation were not significant independent prognostic features.^17,18^ Race emerged as the only significant positive prognostic feature, with white patients experiencing improved overall survival (HR: 0.44). In a pooled analysis of non-TCGA and TCGA data, the impact of race on survival remained significant (Supplement 6C-6D).

We further examined the impact of *EGFR* amplification, *CDKN2A/B* loss, 10q loss, and 22q loss in reducing the median survival for patients with *IDH1/2-*mutant astrocytomas in the non-TCGA cohort. Patients with *EGFR* amplification had a median survival of 37.6 months (range: 2.3-97.1 months), which was less than one-third of the median survival for patients without *EGFR* amplification (120.6 months, range: 0.1-262.2 months, p<0.0001, Supplement 8A). Similarly, patients with homozygous loss of *CDKN2A/B* loss had a median survival of 30.0 months (range: 2.3-167.6 months) compared to 71.6 months if their tumor harbored a heterozygous loss of *CDKN2A/B* (range: 5.8-164.8 months) or 136.8 months with no loss of *CDKN2A/B* (range: 0.1-262.2 months, p<0.0001, Supplement 8B). Patients with 10q loss had a median survival of 43.9 months (range: 0.1-208.8 months), which was less than half the median survival of patients with 10q retained (124.5 months, range: 0.1-262.2 months, p<0.0001, Supplement 8C). Moreover, patients with 22q loss had a median survival of 45.3 months (range: 0.1-128.3 months), compared to 124.7 months for patients with 22q retained (range: 0.1-262.2 months, p<0.001, Supplement 8D). Notably, these negative prognostic molecular features had limited co-occurrence, with only 25.9% of patients with *EGFR* amplification, *CDKN2A/B* loss, 10q loss, and 22q loss having more than one of these features (Supplement 8E).

Prognostic features in non-TCGA *IDH1/2-*mutant 1p/19q-codeleted oligodendrogliomas were rare and no molecular alterations were significant, including *MGMT*-methylation status. High tumor grade was the strongest negative prognostic indicator (HR: 2.71), followed by increasing age per year (HR: 1.06, Fig. 4F). The effect of age and grade were seen in the TCGA cohort as well (Supplement 6E-6F). Of note, while 10q loss did not reach significance, its trend toward being a negative prognostic indicator was consistent with the effect observed in *IDH1/2*-mutant astrocytoma.

### Distribution of Molecular Alterations Changes with Age

Given the prognostic significance of age for glioblastoma and oligodendroglioma, we investigated overall survival in different glioma subtypes across age strata. Patients ≥65 years old with glioblastoma, *IDH1/2*-mutant astrocytoma, or oligodendroglioma had lower median survival compared to patients between 40-64 years and 20-39 years (all p≤0.02, Fig. 5A-C).

**Figure 5:**
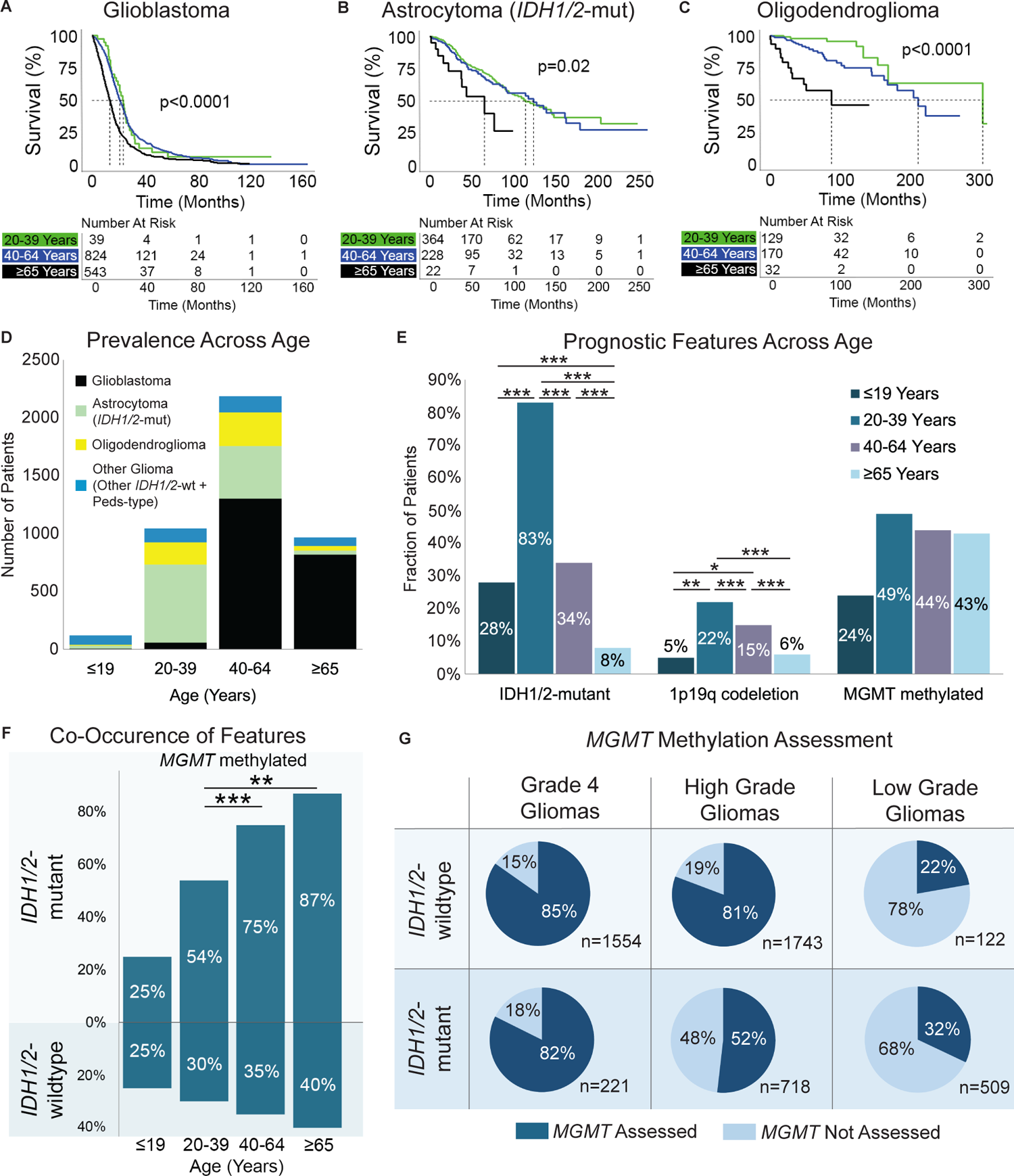
Impact of age on survival and prognostic features. Kaplan-Meier curves for glioma overall survival stratified by age for (A) glioblastoma, (B) *IDH1/2-*mutant astrocytoma, and (C) oligodendroglioma. (D) Prevalence of glioma subtypes across age categories. (E) Prevalence of *IDH1/2*-mutation, *MGMT*-methylation, and chromosome 1p/19q codeletion across age categories. (F) Co-occurrence of *MGMT*-methylation with *IDH1/2* mutation status. (G) Percent of gliomas assayed for *MGMT*-methylation stratified by *IDH1/2* mutation status and tumor grade (high: grades 3-4, low: grade 1-2). p<0.05 (*), p<0.01 (**), p<0.001 (***)

We conducted an epidemiologic survey to examine the prevalence of three molecular signatures with positive prognostic significance across different age groups in glioma: *IDH1/2-*mutation, *MGMT*-methylation and 1p19q codeletion. As expected, across all gliomas, the prevalence of *IDH1/2*-mutant gliomas decreased as patients became older, with only 75 out of 966 patients ≥65 years old having an *IDH1/2-*mutant astrocytoma or oligodendroglioma (Fig. 5D). This decrease reflected the declining prevalence of *IDH1/2*-mutation as patients became older (83% in young adults, 34% in middle aged, and 8% in older adults, X^2^ p<0.001, Fig. 5E). Similarly, the prevalence of 1p19q codeletion showed a decreasing trend with age (22% in young adult, 15% in middle aged, 6% in older adults, X^2^ p<0.001, Fig. 5E). In contrast, the prevalence of *MGMT* promoter methylation was similar across all adult patients, ranging between 43 and 49% across all gliomas (X^2^ p=0.091, Fig. 5E). However, among the few patients over 65 years old with *IDH1/2-*mutation, there was significantly greater co-occurrence of *MGMT*-methylation compared to young adults (Fig. 5F, X^2^ p<0.001). There was no significant difference across age strata for the proportion of patients with co-occurring *IDH1/2*-wildtype status and *MGMT*-methylation (Fig. 5F, X^2^ p=0.154).

Despite the clinical implications of *MGMT* promoter methylation in gliomas, a significant proportion of gliomas were not assessed for *MGMT*-methylation status, especially amongst *IDH1/2*-mutant tumors (Fig. 5G). This was particularly evident among low-grade gliomas, where only 32% of low-grade *IDH1/2*-mutant gliomas were assessed for *MGMT*-methylation status compared to 52% of high-grade and 82% of grade 4 *IDH1/2*-mutant gliomas.

## Discussion

The integration of molecular criteria with histopathological features for glioma classification has significantly advanced neuro-oncology, linking changes in genotype to tumor phenotype and clinical behavior. In our analysis, we provide an updated and contemporary overview of survival estimates across glioma subtypes, dissect pathways involved in glioma tumorigenesis, and characterize important clinical and molecular prognostic features. By directly comparing non-TCGA and TCGA patient cohorts, we emphasize the importance of using updated and clinically representative datasets when developing and validating molecular biomarkers.

Stratification by more contemporary versus the TCGA cohort revealed noteworthy increases in survival rates across various glioma subtypes compared to previous estimates. For instance, patients with glioblastoma in the non-TCGA cohort had a median survival of 19.0 months, exceeding the median survival of 15.0 months observed in previous clinical trials and in the TCGA cohort.^19^ This improvement may be attributed to multiple factors such as the widespread use of chemoradiation, evolution of medical technologies coupled with management of patient complications, and the increase in investigational agents.^20^ However, despite these advances, the median survival of glioblastoma remains relatively low compared other solid and liquid cancers, underscoring the limited progress in reducing the mortality of glioblastoma over the past decade. In comparison, *IDH1/2-*mutant astrocytomas and oligodendrogliomas have seen more significant increases in median survival during the contemporary period: non-TCGA *IDH1/2-*mutant astrocytoma patients had a median survival of 11.4 years while median survival for oligodendrogliomas was 25.6 years. These survival outcomes are more than double what was seen in the TCGA cohort as well as other population wide estimates.^21,22^ The substantial improvements in survival for these two glioma subtypes may reflect the recognition of survival advantage with early surgery or increased utilization of adjuvant radiation and chemotherapy.^23^ In addition, advances in neurosurgical technologies that enable safer and maximal safe tumor resection could contribute to more durable tumor control.^24^ Across glioblastoma, *IDH1/*2-mutant astrocytoma, and oligodendroglioma, heterogeneity in patient populations may have contributed to the differences between non-TCGA and TCGA survival estimates, as non-TCGA sites were often large, urban tertiary care medical centers. Nevertheless, the notable divergence in survival between patients in the non-TCGA versus TCGA cohorts indicates the importance of using contemporary patient profiles when assessing standards of care and novel therapeutics for gliomas.

Through multivariate analysis of a highly powered cohort, we successfully identified both well-established and novel prognostic features that were specific to glioma subtypes. In glioblastoma, several factors were confirmed to be prognostic, including older age, *MGMT*-methylation, receipt of chemotherapy, loss of *CDKN2A/B*, *PIK3CA/3R1*, 19q gain, and *RB1* alteration, which are consistent with previous findings.^4,5,25,26^ However, mutations in *NF1* emerged as a novel negative prognostic marker.^27–29^ Further, we identified 21q loss as a novel positive prognostic marker in glioblastoma. Though the exact molecular mechanism for 21q loss will require further investigation, an in-vitro study of glioblastoma showed significant slowing of cellular proliferation following chromosome 21 loss.^30^ We also demonstrate that heterozygous loss of *CKDN2A/B* conferred a negative impact on overall survival similar to homozygous loss of *CDKN2A/B*, suggesting prognostic value of *CDKN2A/B* in glioblastoma even with incomplete loss.

In the case of *IDH1/2*-mutant astrocytomas, we corroborated several negative prognostic features that were reported in the literature, including alterations in *CCND2*, *PDGFRA*, loss of chromosome 9p, and either homozygous or heterozygous loss of *CDKN2A/B*.^5,31–35^ Additionally, we identified *EGFR* amplification and chromosome 22q loss as novel negative prognostic features. These features have been associated with poor outcome and malignant transformation in other glioma subtypes, especially glioblastoma, but our results demonstrate their significance in defining a subset of more aggressive *IDH1/2*-mutant astrocytomas. Notably, these new prognostic features did not reach statistical significance in the TCGA cohort, highlighting the population heterogeneity between non-TCGA and TCGA cohorts.^36–38^ Moreover, we observed that being of white race emerged as a favorable prognostic indicator for *IDH1/2*-mutant astrocytomas. This finding raises concerns about structural disparities in access to timely surgical and medical care or clinical trial enrollment, which may affect treatment outcomes.^39^

In comparison to glioblastoma and *IDH1/2*-mutant astrocytoma, oligodendroglioma displayed no specific molecular alterations associated with prognosis.^5^ This observation suggests that oligodendrogliomas may be characterized by a more diverse array of genomic alterations correlating with tumor grade, rather than a few dominant recurrent driver mutations dictating higher grade behavior.

Increased patient age is associated with unfavorable prognostic features, as previously demonstrated in smaller cohorts.^40,41^ Older patients were more likely to have higher grade tumors (oligodendroglioma, and astrocytomas) and disadvantageous *PDGFRA* alterations (glioblastoma). Two more favorable features, *IDH1/2*-mutation and 1p19q codeletion decreased as patients aged. Interestingly, the prevalence of *MGMT*-methylation was comparable across age groups. Furthermore, in the limited number of older patients who had *IDH1/2*-mutation, these individuals were significantly more likely to have co-occurring *MGMT*-methylation. This suggests that there is a subset of patients ≥65 years old who may have a biologically more favorable subtype of glioma. However, only 32% of patients with low-grade *IDH1/2-* mutant gliomas and 52% of patients with high-grade *IDH1/2-*mutant gliomas have been assessed for *MGMT*-methylation status. This highlights the need for more widespread *MGMT* molecular profiling to identify patients who may benefit from available therapies.

Correlation analysis and genomic distance measurements revealed the strong association between the accumulation of molecular alterations, histopathologic grade, and patient survival across glioma subtypes. High-grade oligodendrogliomas (grade 3) and *IDH1/2-*mutant astrocytomas (grade 4) showed a higher mutational burden, whereas low-grade oligodendrogliomas (grade 2) and astrocytomas (grades 2/3) consistently showed negative correlations with specific genomic alterations. As a result, grade 3 oligodendrogliomas and grade 4 *IDH1/2-*mutant astrocytomas displayed distinct survival outcomes compared to their lower-grade counterparts. While there were some similarities in their mutational profiles, each glioma subtype demonstrated unique molecular characteristics. Glioblastoma featured several well-known alterations that largely segregated into distinct tumorigenesis pathways. For example, *CDKN2A/B* alterations were associated with alterations in the PI3K pathway but were mostly mutually exclusive from alterations in the p53 and cell cycle pathways, both of which include *CDKN2A/B*. By contrast, *IDH1/2-*mutant astrocytomas had a broader range of co-occurring deleterious alterations across various pathways, including between *CDKN2A/B* and *EGFR*. Despite these associations, grade 4 *IDH1/2*-mutant astrocytomas demonstrated significant genomic distance from glioblastomas, reinforcing their distinct categorization from glioblastoma. In addition, the relative molecular homogeneity and similar survival between grade 2 and grade 3 *IDH1/2-*mutant astrocytomas underscore the lack of well-defined features that can reliably distinguish across these grades. The unique mutational profile for each glioma subtype supports the notion that a molecularly driven classification system for gliomas can enhance precision and improve the correlation between molecular characteristics and clinical behavior.

Several limitations exist in this analysis, many of which are inherent in using large retrospective datasets. Errors in data entry and storage within large repositories may have influenced the clinical and molecular information used in our cohort, despite our efforts to manually update all available institutional data to extend follow-up length and verify molecular and treatment information. Additionally, as the molecular data was collected over two decades, there was heterogeneity in the coverage of certain genes and differences in how mutations were detected. This limited the number of samples with complete molecular data that could be used for survival prognostication. Furthermore, there were constraints on the length of follow-up recorded in clinical outcomes data, particularly for oligodendroglioma patients which may require a considerably long follow-up period to identify important prognostic features accurately. Although we successfully characterized prognostic indicators for glioblastoma, *IDH1/2*-mutant astrocytoma, and oligodendroglioma, we were unable to comprehensively analyze molecular features for the pediatric-type and other *IDH1/2*-wildtype gliomas given the small sample sizes. To better understand the genomic drivers and survival differences for these less common glioma subtypes, a well-powered cohort study is necessary.

Despite these considerations, we believe that this analysis will serve as the framework for further exploration of critical molecular alterations and improved validation of prognostic indicators in gliomas. Given the importance of historical control cohorts for designing and validating clinical trials, our findings serve as a reference for the development of future investigational agents. Insights on glioma molecular profiles and genomic interactions gleaned from this study can further guide recommendations for targeted therapies. The unified resource presented here helps decode the adult glioma landscape, serving as a guidepost for patients and healthcare providers alike.

## Methods

### Patient Cohorts

Patients with clinically and molecularly annotated gliomas were derived from three datasets: 1) Dana-Farber Cancer Institute/Brigham and Women’s Hospital (DFCI/BWH); 2) Project Genomics Evidence Neoplasia Information Exchange (GENIE); and 3) The Cancer Genome Atlas (TCGA). Clinical and molecular data for GENIE and TCGA were downloaded from online repositories while DFCI/BWH data was collected through chart review and institutional next-generation sequencing.^9,42,43^ A full description of data access, molecular profiling, and germline variant filtering can be found in Supplement 9. Age was stratified into four categories: ≤19 years (pediatric); 20-39 years (young adult); 40-64 years (adult); and ≥65 years (older adults). Samples with incomplete genomic profiles or duplicates across cohorts were removed. When multiple glioma sample entries existed per patient, the earliest occurring sample was selected. This study was approved by the Institutional Review Board of the Dana-Farber/Harvard Cancer Center.

### Glioma Classification and Grade

Gliomas were classified into five subgroups based on molecular criteria outlined in the WHO 2021 guidelines and cIMPACT-NOW Updates 1-6: glioblastoma, astrocytoma, oligodendroglioma, pediatric-type gliomas, and other gliomas.^5,6^ Glioblastomas were *isocitrate dehydrogenases 1/2* (*IDH1/2)*-wildtype gliomas with accompanying glioblastoma-associated molecular alterations including *TERT* promoter mutation, *EGFR* copy number amplification, and/or combined whole chromosome 7 gain/chromosome 10 loss (7+/10-). Astrocytomas were *IDH1/2*-mutant gliomas without codeletion of chromosomal arms 1p and 19q. Oligodendrogliomas were *IDH1/2*-mutant gliomas with chromosome 1p19q codeletion. If 1p19q status was not available in an *IDH1/2-*mutant glioma, presence of *ATRX* or *TP53* mutations indicated it was likely an astrocytoma.^44^ 98.8% of mutations in *IDH1/2* were either *IDH1^R^*^132^ or *IDH2^R172^*, while the remainder were non-canonical mutations. Low-grade pediatric-type gliomas were *IDH1/2*-wildtype gliomas with *MAPK* alterations and no glioblastoma-specific alterations. High-grade pediatric-type gliomas were *IDH1/2*-wildtype with *H3K27* or *H3G34* mutation. Finally, other *IDH1/2*-wildtype gliomas included the remaining *IDH1/2*-wildtype and diffuse astrocytic gliomas as well as those labeled “not elsewhere classified (NEC).”^45^

Following molecular reclassification, all glioblastomas were designated as grade 4. Oligodendrogliomas were classified as grade 3 and *IDH1/2*-mutant astrocytomas as grade 4 if they had homozygous deletion of *cyclin-dependent kinase inhibitor 2A* and *2B* (*CDKN2A/B*).^5^ As there are no molecular criteria to distinguish grade 2 from grade 3 astrocytomas, these grades were assigned based on the clinically annotated grade. Grade 1 gliomas when present were also designated using their original annotated grade.

### Molecular Variant Prevalence

Variants of interest were selected if assayed in at least one of the three glioma cohorts. Variant prevalence was calculated based on the total numbers of samples assayed for that gene. To capture a broad set of variants, we selected mutations, copy number variants (CNV), structural variants (SV), or arm-level changes with >1% prevalence. Statistical comparison of molecular alteration prevalence across age groups was performed using Chi-square and pair-wise proportion tests with Holm-Bonferroni correction at a significance level of p<0.05.

### Molecular Correlations and Genomic Distance

Correlations between molecular alterations and clinical variables were examined within glioblastoma, *IDH1/2-*mutant astrocytoma, and oligodendroglioma using the Fisher’s Exact test. *IDH1/2* was excluded as a feature as it was used to define glioma subtypes. Given the multiple genomic alterations explored, we used permutation testing with a Curveball algorithm to generate null molecular alteration matrices which enabled statistical comparison between individual molecular features.^46,47^ Copy number variants and mutations were treated independently to generate the null alteration matrix. Clinical and molecular correlations were significant after corrections for multiple comparisons using the false discovery rate approach with significance q<0.1. Genomic distance within and between glioma subtypes for gene mutations was quantified using the Jaccard similarity index: a Jaccard distance of 1 indicates the greatest genomic difference.^48^ Statistical comparison of Jaccard distances was performed using analysis of variance (ANOVA) with post-hoc Tukey’s test at a significance level of p<0.05.

### Survival Analysis

Overall survival and prognostic features were examined in primary glioblastoma, *IDH1/*2-mutant astrocytoma, and oligodendroglioma for patients ≥20 years old. All patients in the TCGA cohort were diagnosed or operated on in between 1989-2013 while the majority (97.6%) of patients in the non-TCGA cohort (DFCI/BWH and GENIE) were diagnosed or operated on between 2006-2020 (Supplement 10). Date of diagnosis was only used if date of surgery was not available. Kaplan-Meier curves were generated to compare survival between glioma groups, with significance deemed when p<0.05. Upper limits of survival ranges were the time at which all patients were deceased or when last follow-up was completed.

To determine significant prognostic features for overall survival, an initial univariate Cox analysis was separately performed for glioblastoma, *IDH1/*2-mutant astrocytoma, and oligodendroglioma in the non-TCGA cohort. As extent of tumor resection data was not available from the GENIE and TCGA datasets, this was not included as a covariate across all gliomas. Given the documented significance of *MGMT*-methylation on treatment response, glioblastoma samples were selected only if *MGMT*-methylation status was known. For univariate analysis, molecular features were selected as significant if q<0.2 after multiple comparisons correction and prevalence was >3.5% within the glioma type assessed. Features selected as significant on univariate analysis were passed to a multivariate Cox regression model performed for each glioma subtype, along with additional clinical features (patient age, sex, race, receipt of chemotherapy, and tumor grade (if applicable)). A stepwise backwards elimination approach was performed to prevent overfitting and remove any features with undefined confidence intervals. Multivariate features within each glioma subtype were tested on the non-TCGA cohort (DFCI/BWH+GENIE), TCGA cohort, and overall patient cohort. Adjusted features were significant if p<0.05.

## Supporting information

Supplementary Content

## Data Availability

All data produced in the present study are available upon reasonable request to the authors.

## Acknowledgements

We thank the many patients and families that consented to participation in these research studies, along with the DFCI Profile, OncDRS, BWH Center for Advanced Molecular Diagnostics, and BWH Neuropathology staff for assistance with data generation and collection.

## Table Legend

**Table 1:** Cohort. Summary table of surveyed gliomas after molecular classification. Percentages are out of total number of samples included or assayed per variable. Chr: chromosome, n: number.

## Supplementary Legend

**Supplement 1:** Oncoprint showing alterations in genes which were altered in 5% of samples of any of the 5 major glioma subtypes or 4% of the entire study cohort. Additionally, arm-level chromosomal alterations shown if altered in ≥10% of the total cohort, ≥20% of a glioma subtype, or if the sum of the proportion of arms altered across glioma subtypes is ≥20%.

**Supplement 2:** Proportion of gliomas with an affected tumorigenic pathway as defined by select canonical genes in each pathway (as shown in Figure 2): receptor tyrosine kinase (RTK), phosphoinositide-3-kinase (PI3K), mitogen-activated protein kinase (MAPK), p53, and cell cycle.

**Supplement 3:** Jaccard distances quantifying genomic difference between (A) glioblastoma and *IDH1/2*-mutant astrocytoma, (B) glioblastoma and oligodendroglioma, and (C) *IDH1/2*-mutant astrocytoma and oligodendroglioma. p<0.001 (***)

**Supplement 4:** Kaplan-Meier curves for overall survival, stratified by grade, for patients in the non-TCGA cohort with (A) oligodendroglioma, (B) *IDH1/2-*mutant astrocytoma, and (C) other *IDH1/2*-wildtype gliomas. Peds-type: LG: low-grade pediatric-type gliomas, DA/NEC: *IDH1/*2-wildtype diffuse astrocytic gliomas/”Not Elsewhere Classified”, Peds-type: HG: high-grade pediatric-type gliomas.

**Supplement 5:** (A) Kaplan-Meier curves for overall survival in patients with glioblastoma, stratified by *CDKN2A/B* status, demonstrate similar reduction in survival between patients with heterozygous or homozygous *CDKN2A/B* loss versus patients with intact *CDKN2A/B*. (B) Kaplan-Meier curves for overall survival in patients with glioblastoma, stratified by loss or retention of chromosome 21q, show 21q loss positively influences survival. *CDKN2A/B* +/-: heterozygous loss, *CDKN2A/B* -/-: homozygous loss.

**Supplement 6:** Multivariate adjusted hazard ratios and 95% confidence intervals (CI) show differential features for overall survival across the TCGA cohort (A, C, E) versus all patients (B, D, F) for: (A, B) glioblastoma, (C, D) *IDH1/2*-mutant astrocytoma, and (E, F) oligodendroglioma.

**Supplement 7:** Kaplan-Meier curves for overall survival in glioblastoma patients in the TCGA cohort, stratified by methylation status, comparing patients who (A) received versus (B) did not receive chemotherapy.

**Supplement 8:** Patients with *IDH1/2*-mutant astrocytomas stratified by (A) *EGFR* amplification, (B) *CDKN2A/B* homozygous and heterozygous loss, (C) 10q loss, and (D) 22q loss show significantly worse overall survival with each of these prognostic features on Kaplan-Meier curves. (E) Alteration status of *IDH1/2-*mutant astrocytomas with either *EGFR* amplification, *CDKN2A/B* loss, 10q loss, and/or 22q loss, show limited co-occurrence of these four negative prognostic features. *CDKN2A/B* +/-: heterozygous loss, *CDKN2A/B* -/-: homozygous loss.

**Supplement 9:** Methodology for glioma cohort extraction and genomic sequencing.

**Supplement 10:** Stacked bar plot of year of glioma sample collection for non-TCGA and TCGA patients included in analysis of survival and prognostic molecular features. Date of sample collection was inferred from the year of surgery; if this was not available, year of diagnosis was used.

## References

1. Weller, M. et al. Molecular classification of diffuse cerebral WHO grade II/III gliomas using genome- and transcriptome-wide profiling improves stratification of prognostically distinct patient groups. Acta Neuropathol 129, 679–693 (2015).

2. Wiestler, B. et al. Integrated DNA methylation and copy-number profiling identify three clinically and biologically relevant groups of anaplastic glioma. Acta Neuropathol 128, 561–571 (2014).

3. Eckel-Passow, J. E. et al. Glioma Groups Based on 1p/19q, IDH, and TERT Promoter Mutations in Tumors. New England Journal of Medicine 372, 2499–2508 (2015).

4. Kessler, T. et al. Molecular differences in IDH wildtype glioblastoma according to MGMT promoter methylation. Neuro Oncol 20, 367–379 (2018).

5. Louis, D. N. et al. The 2021 WHO Classification of Tumors of the Central Nervous System: a summary. Neuro Oncol 23, 1231–1251 (2021).

6. Louis, D. N. et al. cIMPACT-NOW update 6: new entity and diagnostic principle recommendations of the cIMPACT-Utrecht meeting on future CNS tumor classification and grading. Brain Pathology 30, 844–856 (2020).

7. Ho, V. K. Y. et al. Changing incidence and improved survival of gliomas. Eur J Cancer 50, 2309– 2318 (2014).

8. Weller, M. et al. EANO guidelines on the diagnosis and treatment of diffuse gliomas of adulthood. Nat Rev Clin Oncol 18, 170–186 (2021).

9. Clark, K. et al. The Cancer Imaging Archive (TCIA): Maintaining and Operating a Public Information Repository. J Digit Imaging 26, 1045–1057 (2013).

10. Touat, M. et al. Mechanisms and therapeutic implications of hypermutation in gliomas. Nature 580, 517–523 (2020).

11. Verhaak, R. G. W. et al. Integrated Genomic Analysis Identifies Clinically Relevant Subtypes of Glioblastoma Characterized by Abnormalities in PDGFRA, IDH1, EGFR, and NF1. Cancer Cell 17, 98–110 (2010).

12. Barthel, F. P. et al. Longitudinal molecular trajectories of diffuse glioma in adults. Nature 576, 112–120 (2019).

13. The Cancer Genome Atlas Research Network. Comprehensive, Integrative Genomic Analysis of Diffuse Lower-Grade Gliomas. New England Journal of Medicine 372, 2481–2498 (2015).

14. Ceccarelli, M. et al. Molecular Profiling Reveals Biologically Discrete Subsets and Pathways of Progression in Diffuse Glioma. Cell 164, 550–563 (2016).

15. Aldape, K. et al. Glioma through the looking GLASS: molecular evolution of diffuse gliomas and the Glioma Longitudinal Analysis Consortium. Neuro Oncol 20, 873–884 (2018).

16. Ciriello, G., Cerami, E., Sander, C. & Schultz, N. Mutual exclusivity analysis identifies oncogenic network modules. Genome Res 22, 398–406 (2012).

17. Shirahata, M. et al. Novel, improved grading system(s) for IDH-mutant astrocytic gliomas. Acta Neuropathol 136, 153–166 (2018).

18. Reuss, David. E. Updates on the WHO diagnosis of IDH-mutant glioma. J Neurooncol 162, 461– 469 (2023).

19. Stupp, R. et al. Radiotherapy plus Concomitant and Adjuvant Temozolomide for Glioblastoma. New England Journal of Medicine 352, 987–996 (2005).

20. Thomas, A. A., Brennan, C. W., DeAngelis, L. M. & Omuro, A. M. Emerging Therapies for Glioblastoma. JAMA Neurol 71, 1437 (2014).

21. Dong, X. et al. Survival trends of grade I, II, and III astrocytoma patients and associated clinical practice patterns between 1999 and 2010: A SEER-based analysis. Neurooncol Pract 3, 29–38 (2016).

22. Scheie, D. et al. Prognostic variables in oligodendroglial tumors: a single-institution study of 95 cases. Neuro Oncol 13, 1225–1233 (2011).

23. Hervey-Jumper, S. L. et al. Interactive Effects of Molecular, Therapeutic, and Patient Factors on Outcome of Diffuse Low-Grade Glioma. J Clin Oncol 41, 2029–2042 (2023).

24. Orringer, D. A., Golby, A. & Jolesz, F. Neuronavigation in the surgical management of brain tumors: current and future trends. Expert Rev Med Devices 9, 491–500 (2012).

25. Hegi, M. E. et al. MGMT Gene Silencing and Benefit from Temozolomide in Glioblastoma. New England Journal of Medicine 352, 997–1003 (2005).

26. Weller, M. et al. Molecular Predictors of Progression-Free and Overall Survival in Patients With Newly Diagnosed Glioblastoma: A Prospective Translational Study of the German Glioma Network. Journal of Clinical Oncology 27, 5743–5750 (2009).

27. Brennan, C. W. et al. The Somatic Genomic Landscape of Glioblastoma. Cell 155, 462–477 (2013).

28. Yang, P. H. et al. Multivariate analysis of associations between clinical sequencing and outcome in glioblastoma. Neurooncol Adv 4, (2022).

29. The Cancer Genome Atlas Research Network. Comprehensive genomic characterization defines human glioblastoma genes and core pathways. Nature 455, 1061–1068 (2008).

30. Baskaran, S. et al. Primary glioblastoma cells for precision medicine: a quantitative portrait of genomic (in)stability during the first 30 passages. Neuro Oncol 20, 1080–1091 (2018).

31. Kocakavuk, E. et al. Hemizygous CDKN2A deletion confers worse survival outcomes in IDHmut-noncodel gliomas. Neuro Oncol (2023) doi:10.1093/neuonc/noad095.

32. Shirahata, M. et al. Novel, improved grading system(s) for IDH-mutant astrocytic gliomas. Acta Neuropathol 136, 153–166 (2018).

33. Richardson, T. E. & Walker, J. M. CCND2 amplification is an independent adverse prognostic factor in IDH-mutant lower-grade astrocytoma. Clin Neuropathol 40, 209–214 (2021).

34. Yang, R. R. et al. IDH mutant lower grade (WHO Grades II/III) astrocytomas can be stratified for risk by CDKN2A, CDK4 and PDGFRA copy number alterations. Brain Pathology 30, 541–553 (2020).

35. Roy, D. M. et al. Integrated Genomics for Pinpointing Survival Loci within Arm-Level Somatic Copy Number Alterations. Cancer Cell 29, 737–750 (2016).

36. Wijnenga, M. M. J. et al. Prognostic relevance of mutations and copy number alterations assessed with targeted next generation sequencing in IDH mutant grade II glioma. J Neurooncol 139, 349– 357 (2018).

37. Tesileanu, C. M. S., Vallentgoed, W. R., French, P. J. & van den Bent, M. J. Molecular markers related to patient outcome in patients with IDH-mutant astrocytomas grade 2 to 4: A systematic review. Eur J Cancer 175, 214–223 (2022).

38. Umphlett, M. et al. IDH-mutant astrocytoma with EGFR amplification—Genomic profiling in four cases and review of literature. Neurooncol Adv 4, (2022).

39. Taha, B. et al. Missing diversity in brain tumor trials. Neurooncol Adv 2, (2020).

40. Wiestler, B. et al. Malignant astrocytomas of elderly patients lack favorable molecular markers: an analysis of the NOA-08 study collective. Neuro Oncol 15, 1017–1026 (2013).

41. Krigers, A., Demetz, M., Thomé, C. & Freyschlag, C. F. Age is associated with unfavorable neuropathological and radiological features and poor outcome in patients with WHO grade 2 and 3 gliomas. Sci Rep 11, 17380 (2021).

42. André, F. et al. AACR Project GENIE: Powering Precision Medicine through an International Consortium. Cancer Discov 7, 818–831 (2017).

43. Garcia, E. P. et al. Validation of OncoPanel: A Targeted Next-Generation Sequencing Assay for the Detection of Somatic Variants in Cancer. Arch Pathol Lab Med 141, 751–758 (2017).

44. Brat, D. J. et al. cIMPACT-NOW update 5: recommended grading criteria and terminologies for IDH-mutant astrocytomas. Acta Neuropathol 139, 603–608 (2020).

45. Ellison, D. W. et al. cIMPACT-NOW update 4: diffuse gliomas characterized by MYB, MYBL1, or FGFR1 alterations or BRAFV600E mutation. Acta Neuropathol 137, 683–687 (2019).

46. Bůžková, P., Lumley, T. & Rice, K. Permutation and Parametric Bootstrap Tests for Gene-Gene and Gene-Environment Interactions. Ann Hum Genet 75, 36–45 (2011).

47. Strona, G., Nappo, D., Boccacci, F., Fattorini, S. & San-Miguel-Ayanz, J. A fast and unbiased procedure to randomize ecological binary matrices with fixed row and column totals. Nat Commun 5, 4114 (2014).

48. Bass, J. I. F. et al. Using networks to measure similarity between genes: association index selection. Nat Methods 10, 1169–1176 (2013).

