## Supplementary material for "Molecular Landscape and Contemporary Prognostic Signatures of Gliomas": Table 1

| **Variable** | **Glioblastoma** | **Astrocytoma (*IDH1/2*-mut)** | **Oligodendroglioma** | **Pediatric-type Glioma** | **Other *IDH1/2*-wildtype Gliomas** |
|---|---|---|---|---|---|
| Patients, n | 2195 | 1198 | 531 | 205 | 271 |
| Cohort, n (%) | | | | | |
| TCGA NCI | 325 (14.8) | 266 (22.2) | 164 (30.9) | 5 (2.4) | 12 (4.4) |
| DFCI/BWH | 791 (36.0) | 358 (29.9) | 176 (33.1) | 96 (46.8) | 144 (53.1) |
| Genie (v10) | 1079 (49.2) | 574 (47.9) | 191 (36.0) | 104 (50.7) | 115 (42.4) |
| Sex (Female), n (%) | 886 (40.4) | 483 (40.3) | 250 (47.2) | 95 (46.3) | 115 (42.4) |
| Median Age, years (range) | 61 (6-94) | 36 (7-90) | 43 (13-81) | 27 (1-78) | 51 (0-90) |
| Age (years), n (%) | | | | | |
| ≤19 | 5 (0.2) | 26 (2.3) | 6 (1.1) | 50 (31.1) | 32 (12.5) |
| 20-39 | 58 (2.7) | 673 (56.6) | 193 (36.5) | 69 (42.9) | 51 (20.0) |
| 40-64 | 1301 (59.6) | 455 (38.2) | 290 (54.8) | 30 (18.6) | 111 (43.5) |
| ≥65 | 818 (37.5) | 35 (2.9) | 40 (7.6) | 12 (7.5) | 61 (23.9) |
| Race (White), n (%) | 1918 (93.3) | 1009 (91.9) | 463 (92.6) | 139 (82.2) | 208 (86.7) |
| Histopathologic Diagnosis, n (%) | | | | | |
| Glioblastoma | 1918 (87.4) | 297 (24.8) | 2 (0.4) | 59 (28.8) | 152 (56.1) |
| Astrocytoma | 176 (8.0) | 659 (55.0) | 14 (2.6) | 48 (23.4) | 43 (15.9) |
| Oligodendroglioma | 13 (0.6) | 81 (6.8) | 450 (84.8) | 3 (1.5) | 5 (1.8) |
| Other Gliomas | 88 (4.0) | 161 (13.4) | 65 (12.2) | 95 (46.3) | 71 (26.2) |
| Grade, n (%) | | | | | |
| 1 | - | - | - | 21 (16.7) | 16 (8.0) |
| 2 | - | 266 (25.9) | 240 (51.1) | 9 (7.1) | 12 (6.0) |
| 3 | - | 427 (41.5) | 230 (48.9) | 11 (8.7) | 17 (8.5) |
| 4 | 2195 (100.0) | 334 (32.5) | - | 85 (67.5) | 156 (77.6) |
| Molecular Alterations, n (%) | | | | | |
| *TERT* promoter | 1522 (91.0) | 40 (6.0) | 317 (94.3) | 4 (2.3) | 0 (0) |
| *EGFR* amplification | 1038 (47.5) | 17 (1.7) | 0 (0) | 6 (3.1) | 0 (0) |
| Whole Chr7 Gain/Chr10 Loss | 1231 (57.7) | 8 (0.8) | 1 (0.2) | 2 (1.1) | 0 (0) |
| *CDKN2A/B* hom. del. | 1234 (56.5) | 99 (10.1) | 8 (1.5) | 17 (8.7) | 81 (29.9) |
| *PDGFRA* | 248 (11.3) | 77 (6.4) | 20 (3.8) | 33 (16.3) | 40 (14.8) |
| *PTEN* | 1046 (47.7) | 40 (3.3) | 12 (2.3) | 12 (5.9) | 67 (24.7) |
| *ATRX* | 58 (2.6) | 733 (61.3) | 34 (6.4) | 65 (32.5) | 55 (20.3) |
| *TP53* | 562 (25.6) | 1099 (91.7) | 40 (7.5) | 77 (37.6) | 108 (39.9) |

**Table 1**: **Cohort**. Summary table of surveyed gliomas after molecular classification. Percentages are out of total number of samples included or assayed per variable. Chr: chromosome, n: number.
