## Supplementary Content for "Molecular Landscape and Contemporary Prognostic Signatures of Gliomas"

### Supplement 1 – Glioma Mutational Status

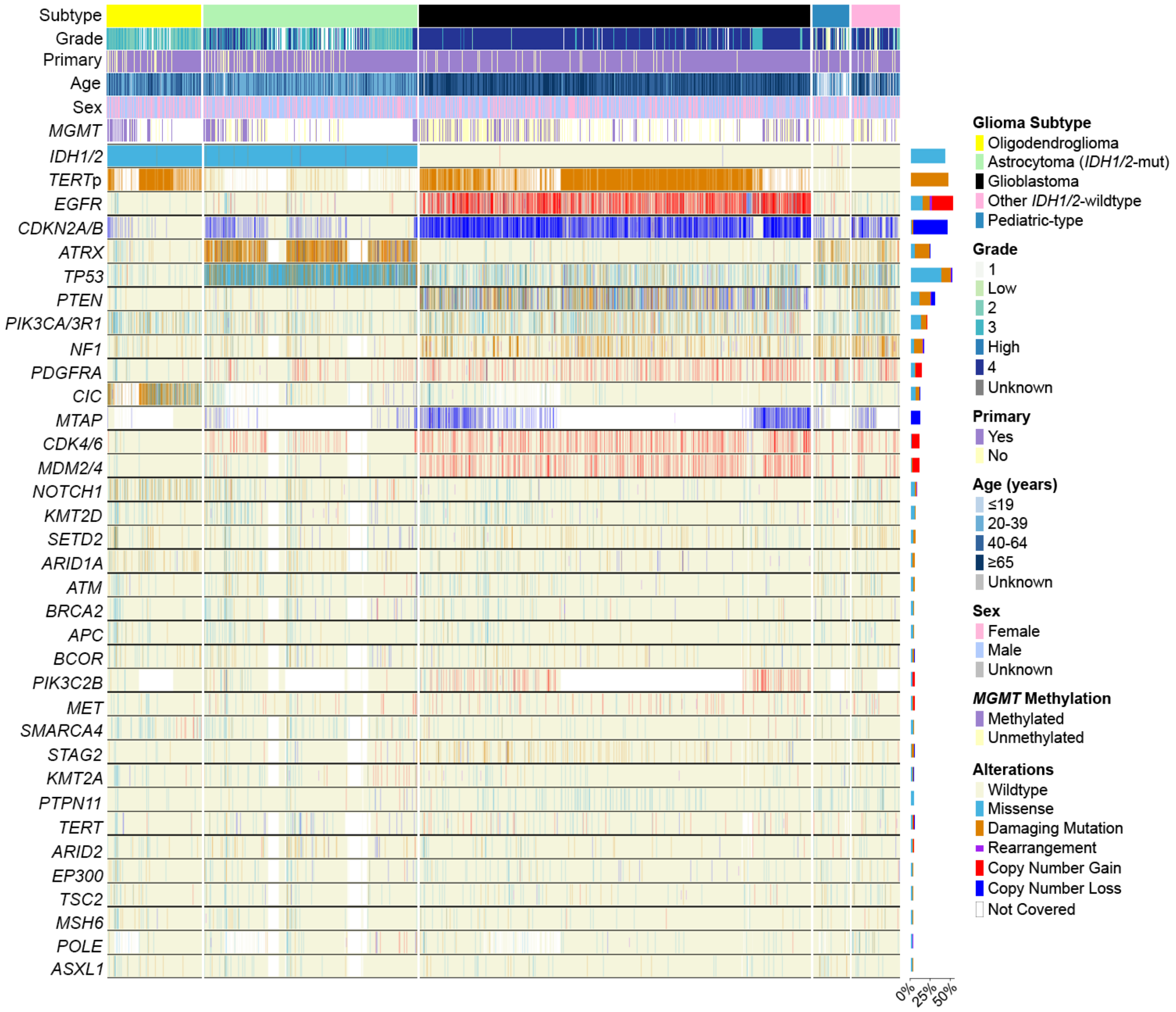

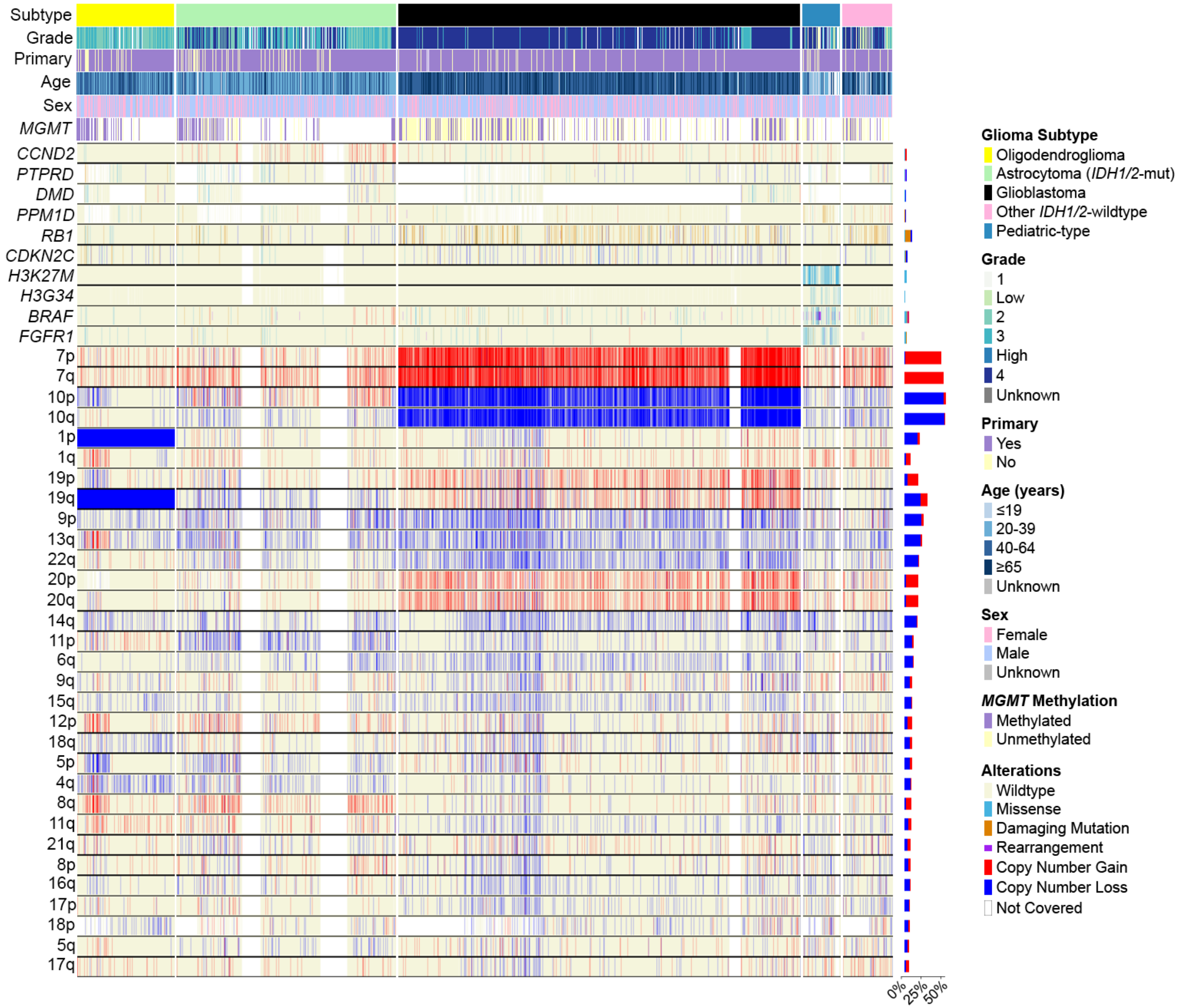

**Supplement 1:** Oncoprint showing alterations in genes which were altered in 5% of samples of any of the 5 major glioma subtypes or 4% of the entire study cohort. Additionally, arm-level chromosomal alterations shown if altered in ≥10% of the total cohort, ≥20% of a glioma subtype, or if the sum of the proportion of arms altered across glioma subtypes is ≥20%.

**Supplement 2 – Pathways Affected Across Glioma Subtypes**

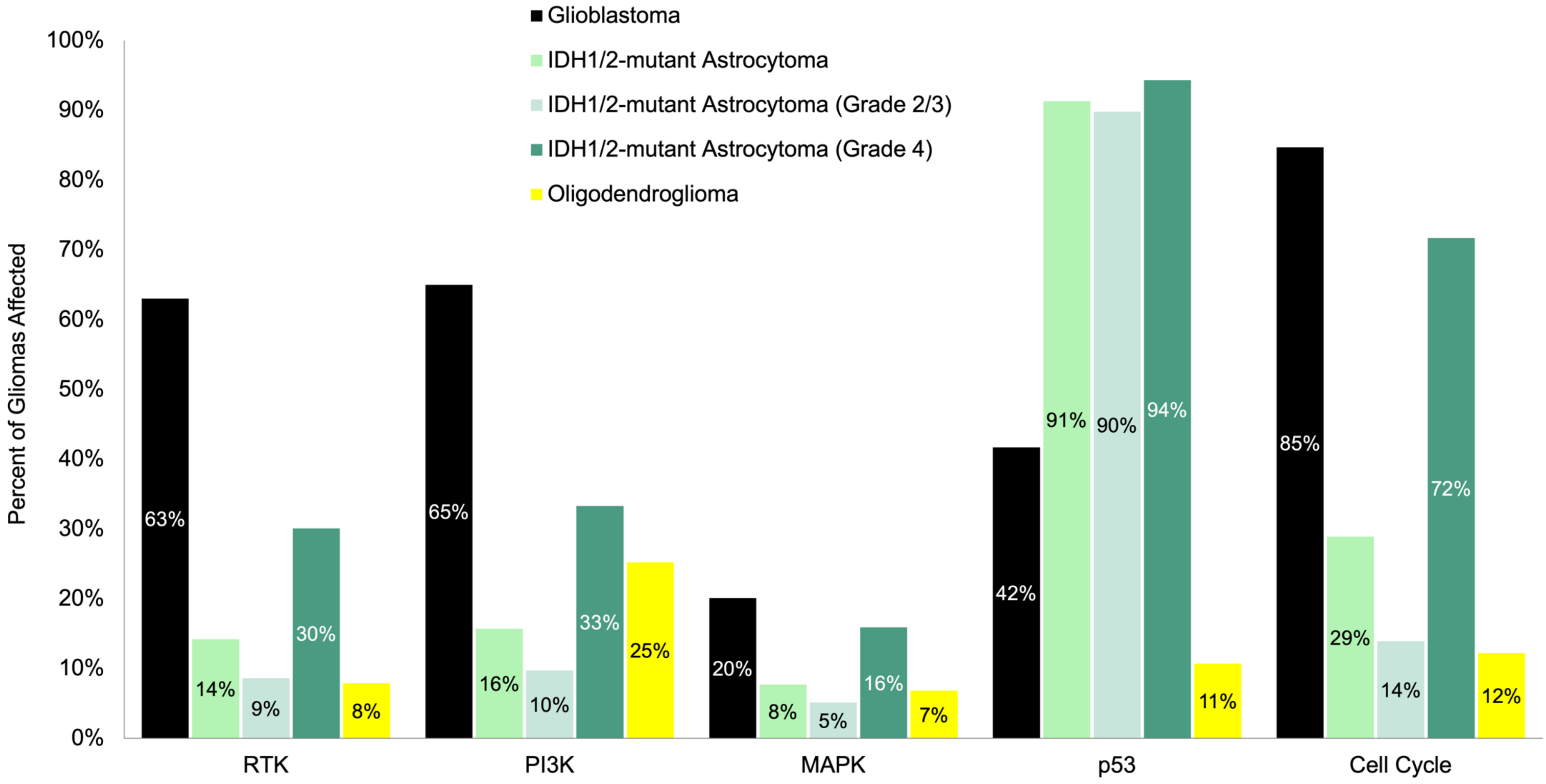

**Supplement 2:** Proportion of gliomas with an affected tumorigenic pathway as defined by select canonical genes in each pathway (as shown in Figure 2): receptor tyrosine kinase (RTK), phosphoinositide-3-kinase (PI3K), mitogen-activated protein kinase (MAPK), p53, and cell cycle.

**Supplement 3 – Genomic Distances across Glioma Subtypes**

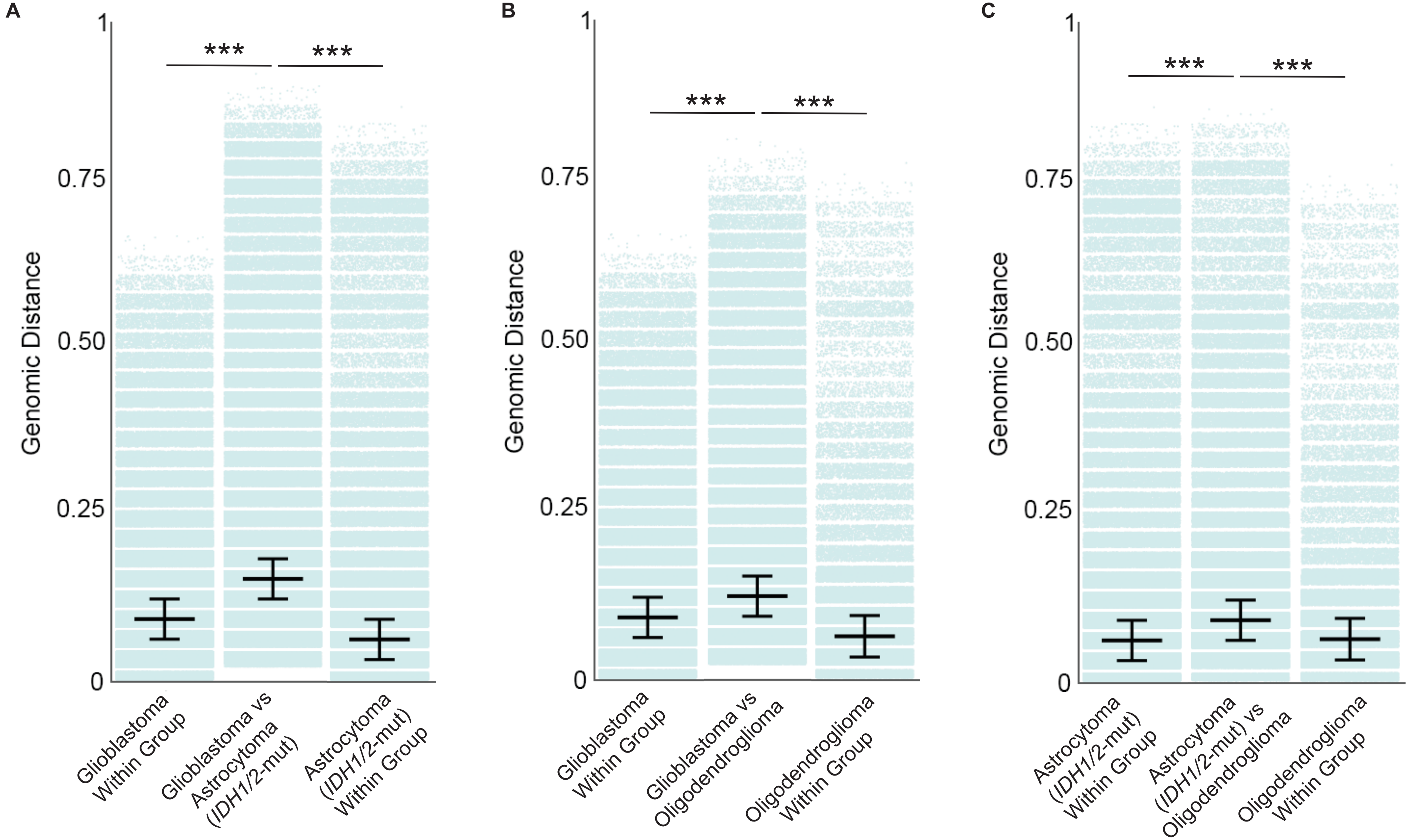

**Supplement 3:** Jaccard distances quantifying genomic difference between (A) glioblastoma and *IDH1/2*-mutant astrocytoma, (B) glioblastoma and oligodendroglioma, and (C) *IDH1/2*-mutant astrocytoma and oligodendroglioma. p<0.001 (***)

**Supplement 4 – Glioma Survival by Grade: Non-TCGA Cohort**

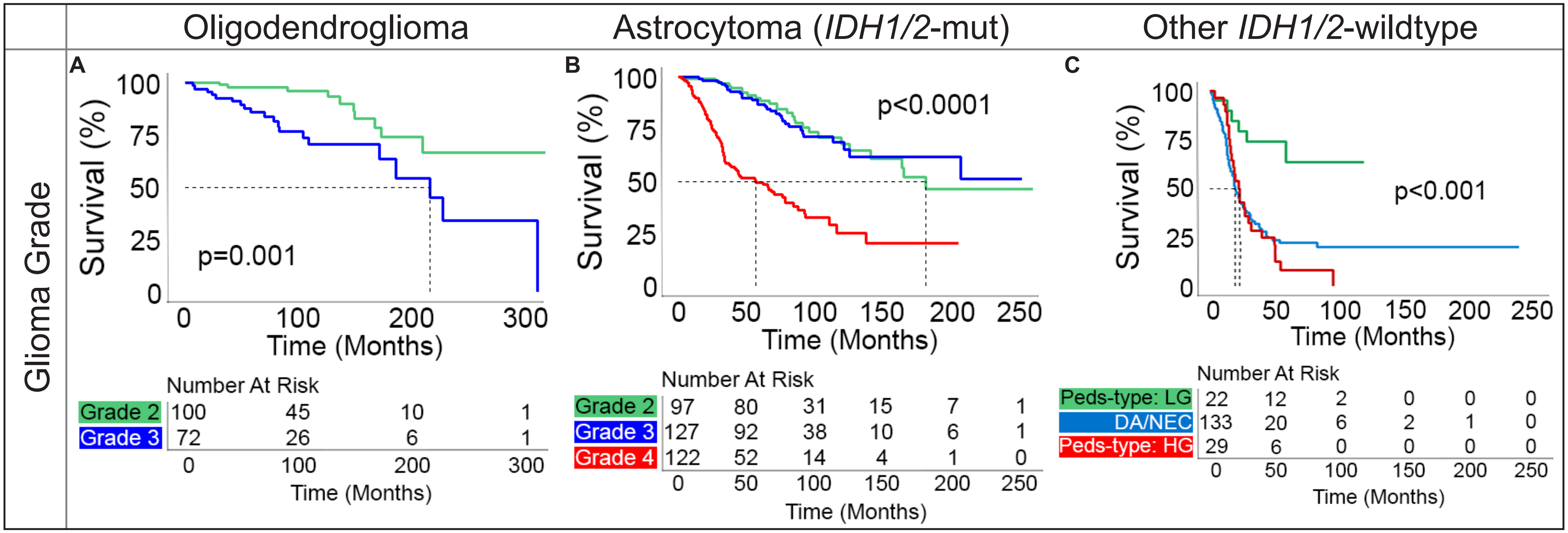

**Supplement 4:** Kaplan-Meier curves for overall survival, stratified by grade, for patients in the non-TCGA cohort with (A) oligodendroglioma, (B) *IDH1/2*-mutant astrocytoma, and (C) other *IDH1/2*-wildtype gliomas. Peds-type: LG: low-grade pediatric-type gliomas, DA/NEC: *IDH1/2*-wildtype diffuse astrocytic gliomas/"Not Elsewhere Classified", Peds-type: HG: high-grade pediatric-type gliomas.

**Supplement 5 – *CDKN2A/B* and Chromosome 21q Loss in Glioblastoma**

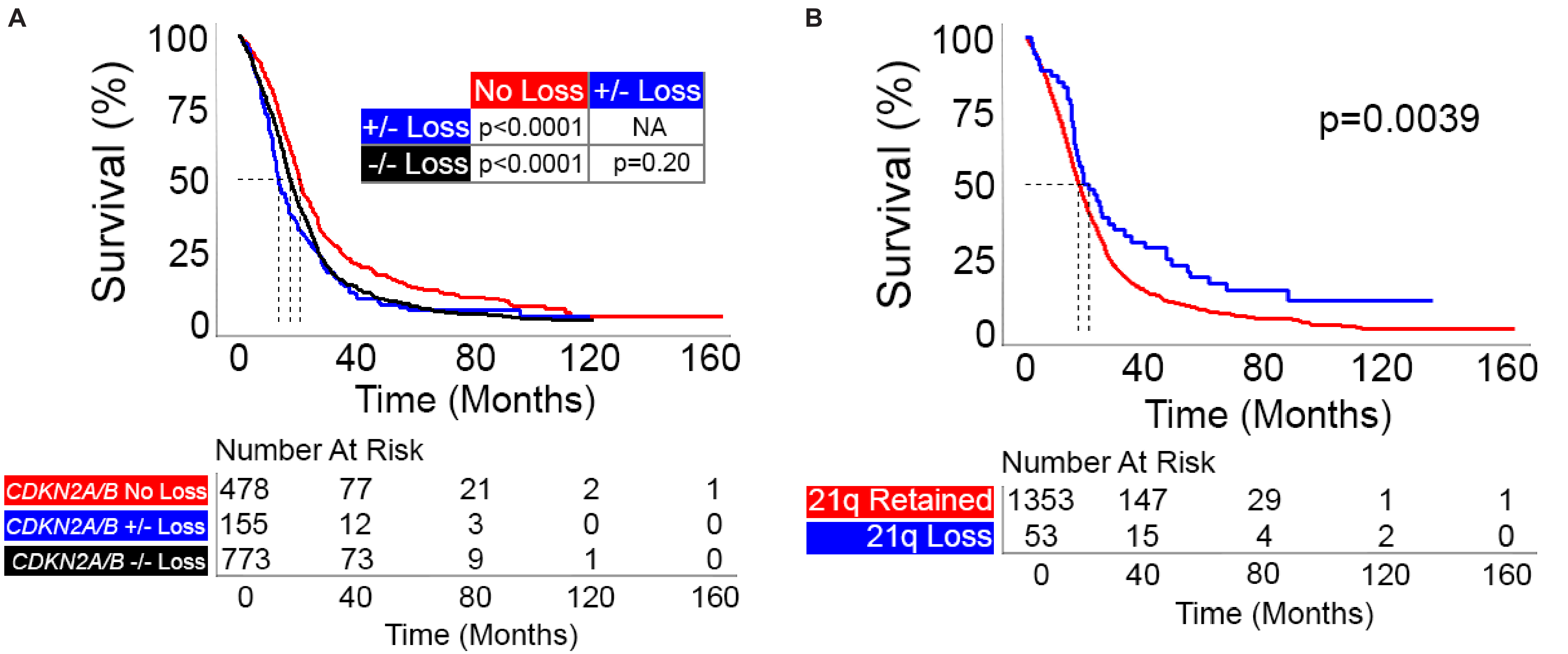

**Supplement 5:** (A) Kaplan-Meier curves for overall survival in patients with glioblastoma, stratified by *CDKN2A/B* status, demonstrate similar reduction in survival between patients with heterozygous or homozygous *CDKN2A/B* loss versus patients with intact *CDKN2A/B*. (B) Kaplan-Meier curves for overall survival in patients with glioblastoma, stratified by loss or retention of chromosome 21q, show 21q loss positively influences survival. *CDKN2A/B* +/-: heterozygous loss, *CDKN2A/B* -/-: homozygous loss.

**Supplement 6 – Multivariate Adjusted Prognostic Features: TCGA Cohort and All Patients**

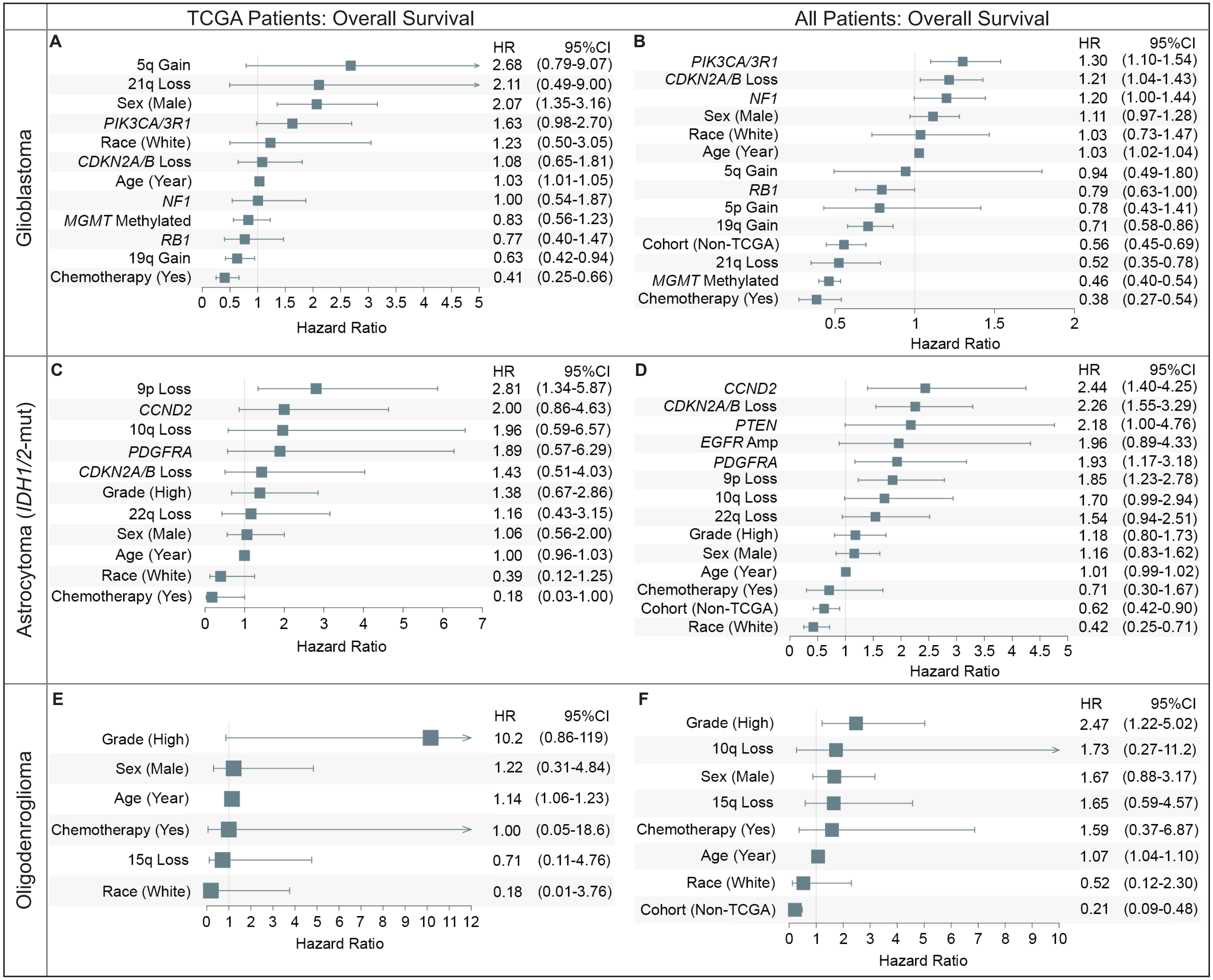

**Supplement 6:** Multivariate adjusted hazard ratios and 95% confidence intervals (CI) show differential features for overall survival across the TCGA cohort (A, C, E) versus all patients (B, D, F) for: (A, B) glioblastoma, (C, D) *IDH1/2*-mutant astrocytoma, and (E, F) oligodendroglioma.

**Supplement 7 – Treatment Status and *MGMT*-Methylation in Glioblastoma Patients from TCGA Cohort**

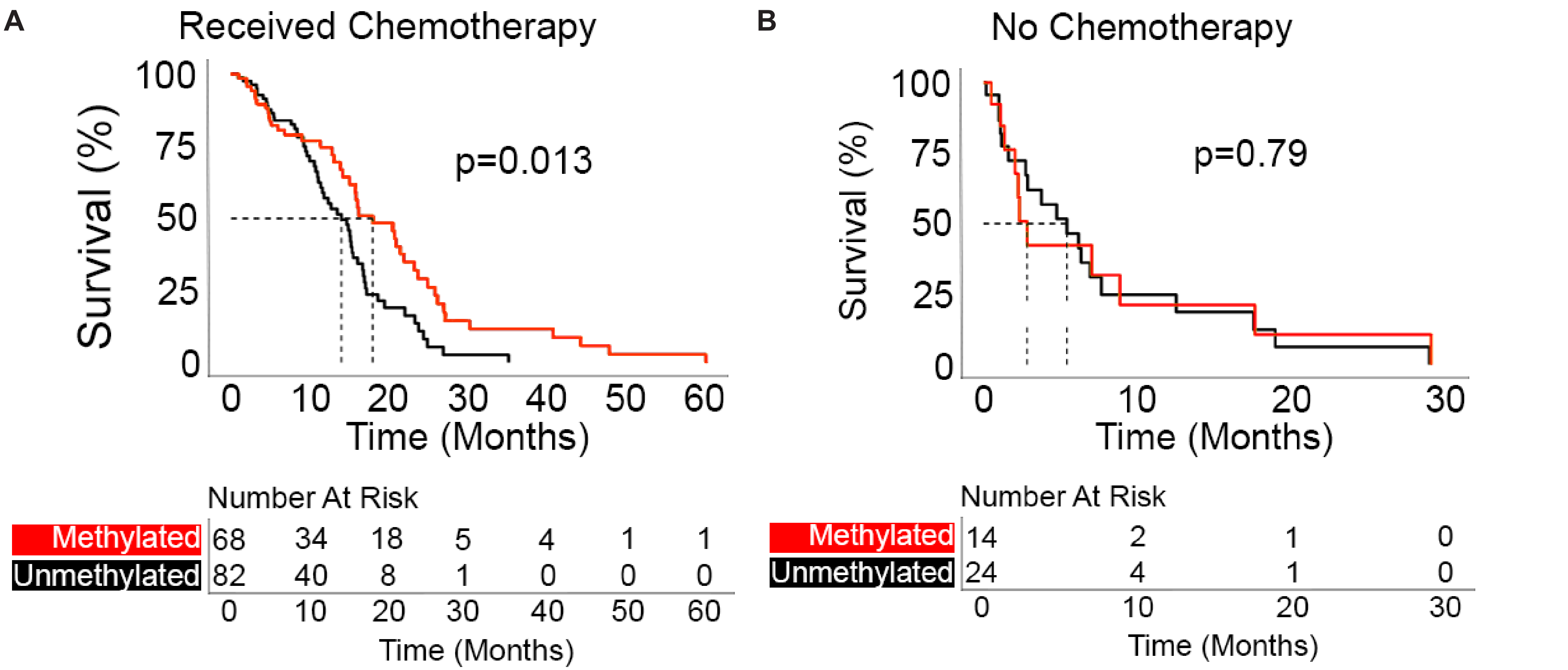

**Supplement 7:** Kaplan-Meier curves for overall survival in glioblastoma patients in the TCGA cohort, stratified by methylation status, comparing patients who (A) received versus (B) did not receive chemotherapy.

### Supplement 8 – Prognostic Signatures in *IDH1/2*-mutant Astrocytoma

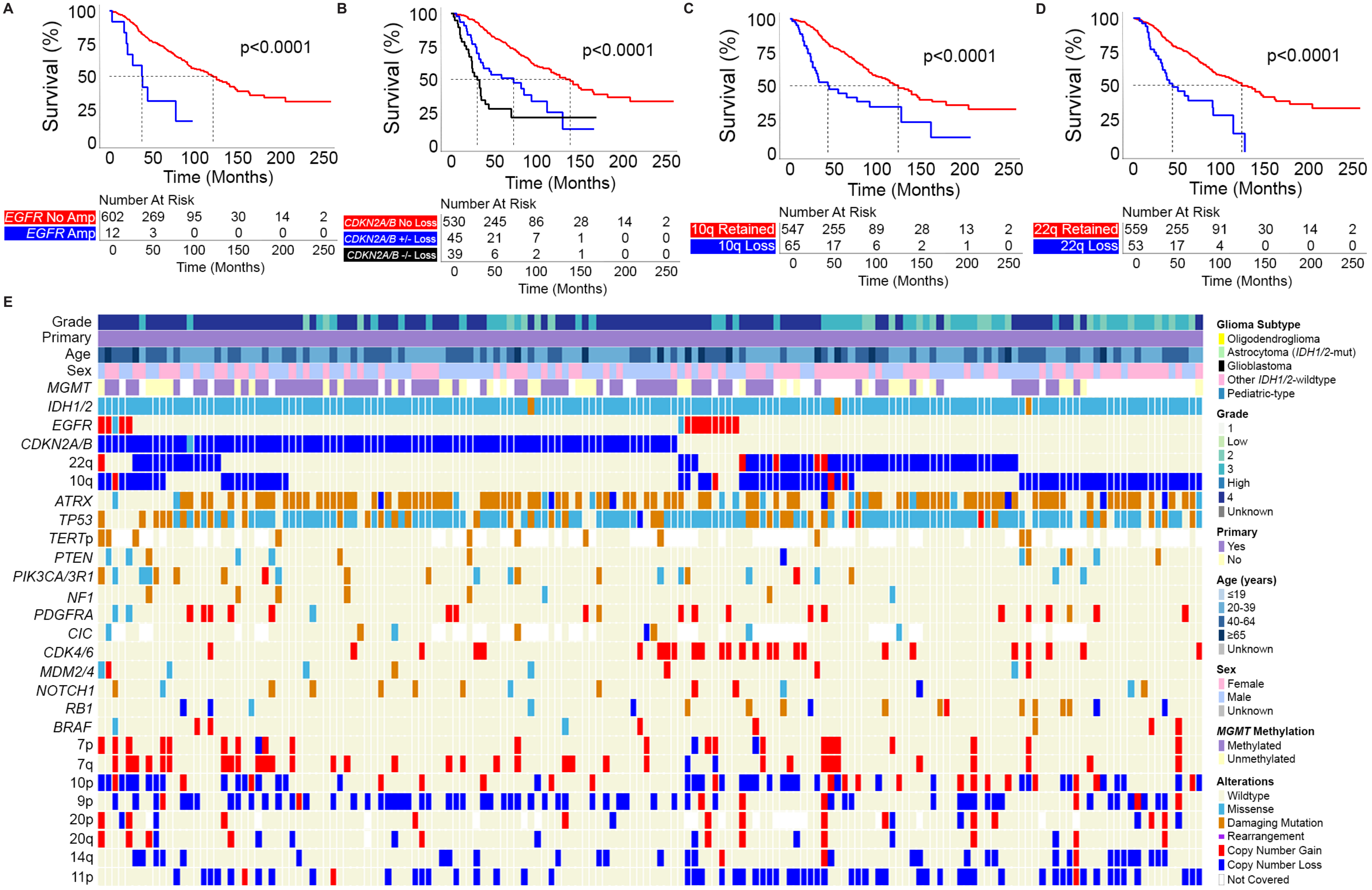

**Supplement 8:** Patients with *IDH1/2*-mutant astrocytomas stratified by (A) *EGFR* amplification, (B) *CDKN2A/B* homozygous and heterozygous loss, (C) 10q loss, and (D) 22q loss show significantly worse overall survival with each of these prognostic features on Kaplan-Meier curves. (E) Alteration status of *IDH1/2*-mutant astrocytomas with either *EGFR* amplification, *CDKN2A/B* loss, 10q loss, and/or 22q loss, show limited co-occurrence of these four negative prognostic features. *CDKN2A/B* +/-: heterozygous loss, *CDKN2A/B* -/-: homozygous loss.

### Supplement 9 – Cohort Extraction and Sequencing

TCGA:

Clinical and molecular data for 1,020 gliomas collected between 1989-2013 were downloaded from the National Cancer Institute (NCI) Genomic Data Commons (TCGA-Low Grade Glioma and TCGA-Glioblastoma datasets, https://gdc.cancer.gov). All TCGA data provided mutational status (spanning 846 genes), copy number variants (CNV), and structural variants (SV) for included glioma samples, which was determined through whole exome sequencing. Germline variants were filtered out from samples using tumor-matched sequencing data. Chromosomal arm level calls were generated through the Genomic Identification of Significant Targets in Cancer (GISTIC) module.

DFCI/BWH:

2,090 glioma samples from DFCI/BWH were assessed between 1993-2020. Clinical data and *MGMT* methylation status were extracted through retrospective review of the electronic medical record. Patient follow-up was collected through July 1, 2023. Through the DFCI-Profile initiative, targeted next-generation sequencing (OncoPanel) was performed at the Center for Advanced Molecular Diagnostics at Brigham and Women's Hospital. OncoPanel results include mutational status, CNVs, and SVs. The number of genes assayed varied based on OncoPanel version, spanning from 277 (OncoPanel V1), 302 (OncoPanel V2), or 477 genes (OncoPanel V3).

As OncoPanel sequencing was performed on glioma samples without matched normal DNA from patients, an internally developed pipeline was applied for germline variant filtering. Variants were first filtered out if their allele frequency was >0.1% in the Genome Aggregation Database (https://gnomad.broadinstitute.org) or if annotated as benign in the NIH ClinVar database (https://ncbi.nlm.nih.gov/clinvar). In the second step, these filtered variants were cross-referenced and added back if present in the Catalogue of Somatic Mutations in Cancer (COSMIC, https://cancer.sanger.ac.uk/cosmic). Chromosomal arm-level calls were generated using the Arm-level Copy-number Events in Targeted Sequencing (ASCETS, Spurr et al., 2020: https://doi.org/10.1093/bioinformatics/btaa980) platform developed at DFCI. Chromosome arms were considered amplified or deleted if more than 70% of the chromosome arm was altered.

GENIE (v10):

Clinical and molecular data for 7,271 gliomas were downloaded from Synapse (17 institutions contributing glioma specific data, https://synapse.org/genie). Samples from DFCI/BWH were filtered out. A total of 32 distinct targeted-sequencing gene panels were applied across the glioma samples included in the filtered GENIE repository. 12 of these gene panels provided CNVs and 9 provided SVs. *IDH1/2*-wildtype and *H3*-wildtype gliomas were excluded if sufficient molecular data was not present to determine if it was an *IDH1/2*-wildtype glioblastoma. Applying this restriction resulted in glioma samples from 1986-2020, which were surveyed on 30 distinct gene panels, representing 45-760 genes. Germline variants were filtered out by an internal pipeline at the American Association of Cancer Research as well as by the institutions contributing data to GENIE. Chromosomal arm level calls were generated using ASCETS using the same sample amplification and deletion definition as DFCI/BWH. *MGMT* methylation data was reported and available only for a subset of GENIE gliomas, specifically those collected and reported by Memorial Sloan Kettering Cancer Center. Patient follow-up was updated using GENIE v13 released in April 2023 on Synapse.

**Supplement 10 – Histogram of Year of Glioma Sample Collection**

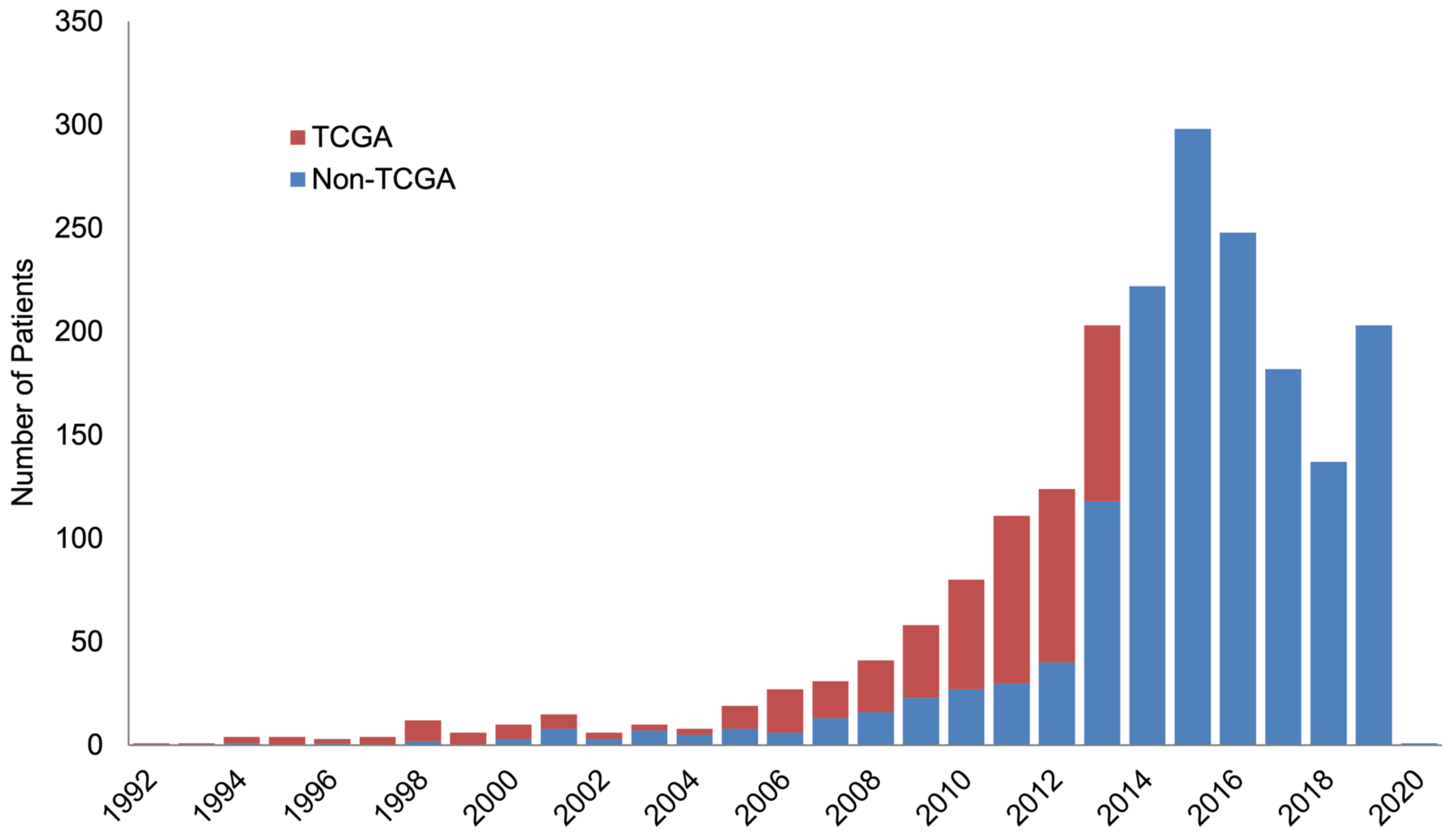

**Supplement 10:** Stacked bar plot of year of glioma sample collection for non-TCGA and TCGA patients included in analysis of survival and prognostic molecular features. Date of sample collection was inferred from the year of surgery; if this was not available, year of diagnosis was used.
